# The construction of admission stroke severity centiles by time from symptom onset to assessment

**DOI:** 10.1101/2025.11.08.25339811

**Authors:** Zewen Lu, Halvor Næss, Matthew Gittins, Amit K Kishore, Craig J Smith, Andy Vail

**Author notes:** **Corresponding author** Zewen Lu, Jean McFarlane Building, University of Manchester, Oxford Road, Manchester M13 9PL. **Statements**. **Author contributions** Zewen Lu: Conceptualisation; Methodology; Study design and data analysis; Writing-original draft. Writing-review & editing. Halvor Næss: Data management and acquisition; Ethical approval; Writing-review & editing. Matthew Gittins, Amit K Kishore, Craig J Smith and Andy Vail: Conceptualisation; Methodology; Supervision; Writing-review & editing. **Funding** None. **Ethical considerations** This study was approved by the Regional Ethics Committee for Medical and Health Research Ethics in Western Norway (approval no. 2012/1483 Norstroke). **Data Availability** All data produced in the present study are available upon reasonable request to the authors.

## Abstract

Centiles provide a framework for interpreting individual measurements by comparing them to a reference distribution. While centile-based approaches have been widely used in fields like perinatology, their potential to stroke severity has not been explored. This study aimed to construct centiles of admission stroke severity by time from symptom onset to assessment, creating a reference that captures the temporal dynamics of stroke severity.

We evaluated three methods—Generalised Additive Models for Location, Shape, and Scale (GAMLSS), Quantile Regression (QR), and the Healy Rasbash Yang (HRY) method—to account for the distinctive characteristics of ischaemic stroke and intracerebral haemorrhage. The parametric GAMLSS approach allowed for non-negative integer values suitable for the discrete nature of stroke severity scores. QR and the HRY method, as nonparametric approaches, provided flexibility in smoothing centiles.

We are the first to construct centile curves for admission stroke severity, offering a novel and meaningful approach for interpreting stroke severity at admission. These centiles have the potential to enhance clinical decision-making and research applications, such as defining stroke severity categories, improving patient monitoring protocols, refining clinical trial design and enhancing stroke prognostication.

## Introduction

Centiles, or percentiles, represent points in a distribution of a given measurement that divides the dataset into 100 equal parts ^1,2^. Centiles provide a way to understand the position of an individual value relative to the entire population. They are commonly used in clinical and healthcare settings to compare an individual’s measurement to a reference population, primarily when associated with a covariate, such as time ^3^.

Reference centile charts are constructed and modelled as a series of smoothed curves to show how measurement changes when plotting against time ^4^. Such charts are widely used in perinatology. For example, when assessing a baby’s birth weight, a two-kilogram newborn may be considered normal for 28 weeks of gestation but too small for 36 weeks. Comparing their birth weight with the birth weight of a large population of babies at a similar gestational age can help identify those outside the normal range who may require monitoring ^5,6^.

The application of reference centiles is not yet widely extended beyond anthropometric measurement to assess human growth and development. However, reference centiles are potentially helpful in addressing a similar statistical issue in the stroke field.

Stroke severity, measured on arrival to the hospital, is widely used in clinical practice to inform urgent treatments and prognosis. Just as in perinatology (e.g., assessing birth weight against gestational age), stroke severity can vary depending on some covariates, including time since symptom onset ^7,8^. Stroke symptoms appear and progress distinctively among patients, which reflects the dynamic nature of the underlying pathophysiology ^9^. This means that the severity of symptoms observed at the time of admission represents just a single realisation within an ongoing, dynamic process. Despite this, ‘admission severity’ is often treated as a fixed covariate in both clinical practice and research. Clinically, it serves as a critical factor guiding treatment decisions, such as the appropriateness of thrombolysis or thrombectomy ^10^. In research, admission severity is frequently used as a criterion for study eligibility or as a baseline measure in prognostic modelling ^11,12^. This static interpretation risks oversimplifying a highly temporal and evolving phenomenon, potentially limiting its utility in capturing the nuance in stroke progression.

Also as in perinatology, the timing of realisation of this dynamic process may itself be status-dependent. Just as an intervention to deliver by Caesarean section or induction may be prompted by fetal size, hospital admission for stroke may be accelerated by more severe symptoms. Constructing centiles allows us to compare an individual patient’s stroke severity on admission with others of similar admission time, providing a reference point to assess whether their severity is typical or atypical for their condition and time since symptom onset. To our knowledge, we are the first to construct centiles for stroke severity in the context of the time from symptom onset to assessment. We evaluated and applied different statistical methods to ensure the feasibility and robustness of the centile construction, acknowledging the need for adjustments to existing approaches. Additionally, given the differences in pathology, progression, and severity between ischaemic stroke (IS) and intracerebral haemorrhage (ICH), we modelled admission severity centiles separately for each stroke type 13,14.

## Methods

### Method selection

In our exploration of modelling admission stroke severity centiles, we identified several potential methodologies that could be suitable ^15,16^. Rather than attempting to identify the single best method, we sought to evaluate the feasibility of developing time-adjusted admission stroke severity centiles using various approaches. We considered several critical factors that influenced our method selection:

- Treatment of Time: Whether the method treats time as a continuous or categorical variable. We only consider methods handling time as a continuous variable.
- Centile Estimation: Whether the centiles are estimated separately or jointly. We try to cover both in this study for illustration and comparison.
- Distributional Assumptions: whether there is a need for any type of distributional assumptions regarding admission stroke severity.
- Curve-Fitting Techniques: We only consider methods with curve-fitting methods applied.
- Fit Assessment: The ability to assess the fit of the model to the data. We prefer methods with a clear evaluation framework.
- Practicality: We favoured approaches commonly used in applied medical research, emphasising ease of implementation, methodological clarity, and the availability of coding resources.

Based on these criteria, we selected three distinct methods, including Generalized Additive Models for Location, Shape, and Scale (GAMLSS) ^17,18,19^, Quantile Regression (QR) ^20,21,22,^ and the HRY method ^23,24^ to provide a balanced approach, incorporating both parametric and nonparametric perspectives. The list of potential methods and the selection process is presented in Appendix F Table 7. By considering a method with distributional assumptions (GAMLSS), one without (QR), and a fully nonparametric approach (HRY), we assessed the feasibility of developing time-adjusted admission stroke severity centiles across different modelling frameworks. GAMLSS and QR are two of the most recognised methods for centile estimation, and the available R packages are easy to implement. The HRY method has no R package, but the coding and computation are relatively straightforward.

### Stroke severity

The National Institutes of Health Stroke Scale (NIHSS) is the most widely used tool for assessing stroke severity in acute settings ^25^. Developed to standardise the evaluation of neurological deficits, the NIHSS evaluates various domains of neurological function through 15 items, each scored on a predefined scale (Table 1). These items measure key functions such as consciousness, motor abilities, language, sensory perception, and coordination. The scores are summed to produce a total NIHSS score ranging from 0 to 42, where higher scores indicate more severe impairment ^26,27^. A score of zero represents the absence of detectable neurological deficits, while higher values reflect increasing severity.

**Table 1.** Scoring Structure of the NIH Stroke Scale (NIHSS)

| No. | NIHSS item | Score | Description |
| --- | --- | --- | --- |
| 1 | Level of consciousness (LOC) | 0–3 | Alertness and responsiveness |
| 2 | LOC questions | 0–2 | Ability to answer questions |
| 3 | LOC commands | 0–2 | Ability to follow simple commands |
| 4 | Best gaze | 0–2 | Horizontal eye movements |
| 5 | Visual fields | 0–3 | Visual field deficits |
| 6 | Facial palsy | 0–3 | Facial muscle movement |
| 7 | Motor arm (left) | 0–4 | Arm strength and movement (left side) |
| 8 | Motor arm (right) | 0–4 | Arm strength and movement (right side) |
| 9 | Motor leg (left) | 0–4 | Leg strength and movement (left side) |
| 10 | Motor leg (right) | 0–4 | Leg strength and movement (right side) |
| 11 | Limb ataxia | 0–2 | Coordination of limb movements |
| 12 | Sensory | 0–2 | Ability to perceive sensory stimuli |
| 13 | Best language | 0–3 | Speech and language comprehension |
| 14 | Dysarthria | 0–2 | Speech clarity |
| 15 | Extinction and inattention | 0–2 | Neglect of stimuli on one side |

### Parametric method with distribution assumption: GAMLSS

#### Model specification

Let outcome Y_i_ represent the admission stroke severity at time t_i_ for patient i where i = 1,…,n). Here, t_i_ denotes the interval from the time of first known symptom onset to the time of admission severity assessment. Assume D represents the chosen distribution of the response variable Y_i_, the conditional distribution of Y_i_ on t_i_ is:

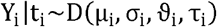

Where µ_i_ is the location (mean) parameter, σ_i_ is the scale(dispersion) parameter, ϑ _i_ is the shape (skewness) parameter, and τ_i_ is the shape (kurtosis) parameter.

Let g_k_ (^*^),where k = 1,2,3,4 to be a known monotonic link function relating the distribution parameters to the explanatory variable t_i_ through an additive model given by

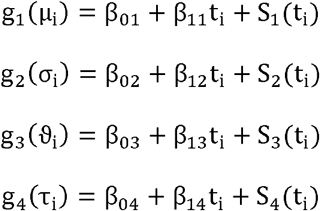

Where S_k_ (t_i_) is the non-linear parametric smoothing function of the explanatory variable time t to affect the location, scale and shape of the fitted conditional response distribution.

### Distributional assumptions

Given the empirical distribution of admission stroke severity in Figure 1, there are a few considerations in selecting the distributional assumptions.

- Continuous or discrete: Understanding the nature of NIHSS as a count-like variable is crucial for modelling stroke severity. The individual items are often treated as discrete integers, resulting in an overall score that reflects both the presence and degree of impairment across multiple domains, as shown in Table 1.
- Overdispersion: Overdispersion exists as the sample variance is much larger than the sample mean for admission NIHSS score of both IS 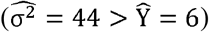 and ICH patients 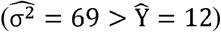.
- Skewness: The distribution of admission NIHSS is positively skewed, especially for IS patients in the first three hours. We binned admission NIHSS by each hour and calculated the skewness for the sample distribution, the skewness also varied over time.
- Zero values: An NIHSS score of zero typically represents the absence of any observable neurological deficits and does not mean the absence of a stroke ^28^. Zero-inflated distributions are not applicable here as zero values in the data are not thought to arise from a different process than the non-zero values ^29^. The zero values also make it challenging to fit most asymmetric continuous distributions as they require Y >0.

**Figure 1.**
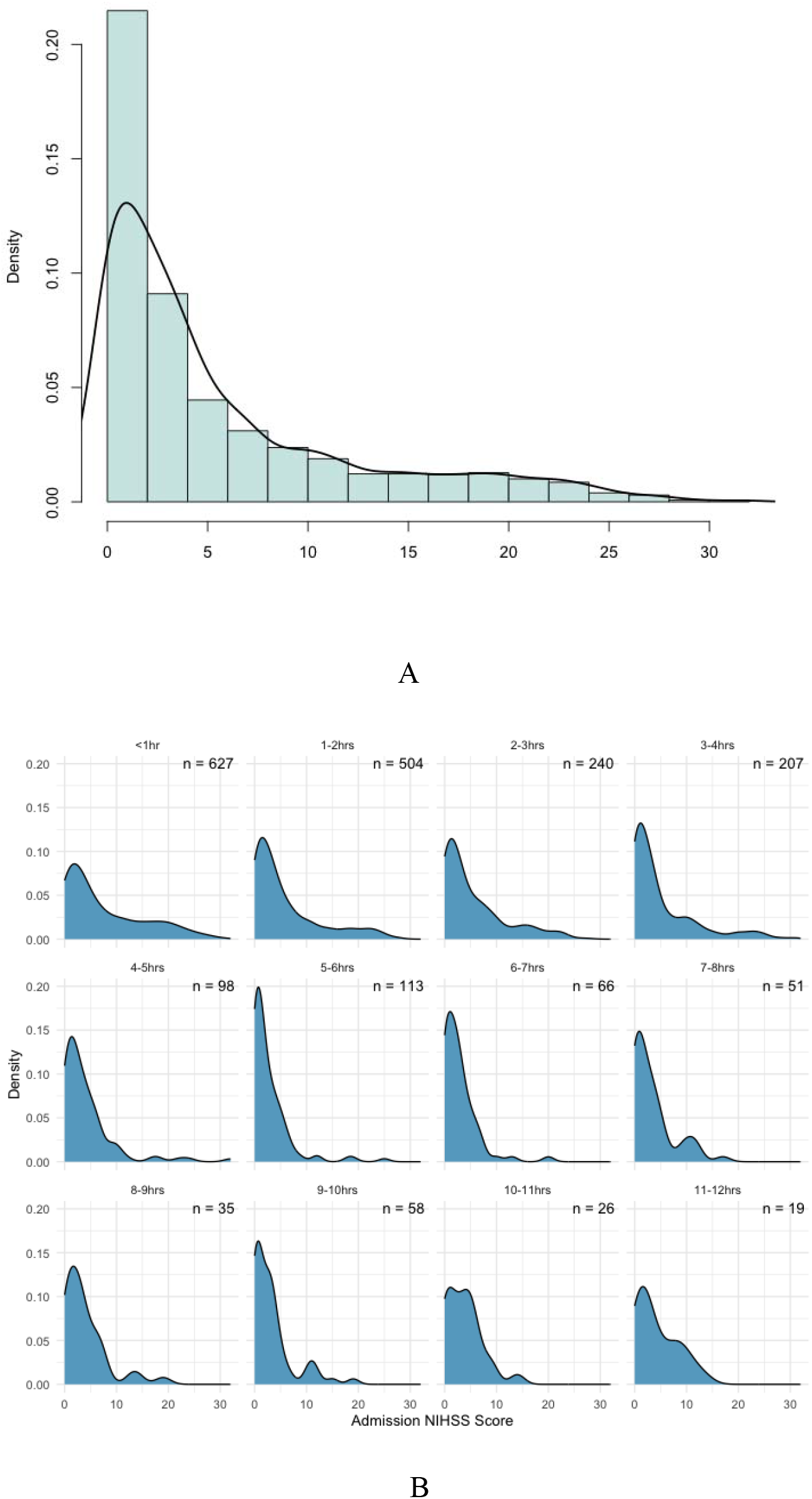
(A) Frequency distribution of admission NIHSS with a kernel density estimate (KDE) of the variable shown as a fitted line (B) Frequency distribution of admission NIHSS stratified by time from symptom onset to assessment in each hour

We therefore chose the negative binomial distribution to represent admission NIHSS containing discrete counts, zero values, overdispersion and skewness. Y|t∼NBI (µ,σ) has mean µ and variance µ+ σµ^2^ as functions of t to vary over time in the GAMLSS model. Negative binomial distribution is a two-parameter distribution. Although the skewness is a function of µ and σ and expected to change over time, there are no explicit β coefficient associated with the shape parameter (skewness) to estimate in the additive models. We, therefore, also tried to fit three-parameter distributions such as beta negative binomial and Delaporte that consider the changes in skewness over time.

### Centile estimation

It is of interest to determine a stroke patient’s position relative to the overall severity distribution of patients admitted at the same time since symptom onset. To achieve this, we modelled the conditional distribution F (Y|t) and estimated the parameters of the fitted model using the Rigby and Stasinopoulos (RS) algorithm by maximising the penalised log-likelihood function ^17^. The 100pth percentile of this distribution, q_p_ (x) = F^-1^ (p|t). defines the admission severity at time t.

We estimated a symmetric subgroup of percentile curves, such as the 10th, 20th, 30th, 40th, 50th, 60th, 70th, 80th, and 90th percentiles. These percentile curves allow for the identification of patients with unusual severity levels, as they highlight measurements that lie in the tails of the appropriate reference distribution.

### Parametric method without distributional assumption: QR

Whereas linear regression aims to estimate the mean, in a classic quantile regression, a pre-specified *p* quantiles (or 100pth percentiles) are estimated directly and separately without any distributional assumptions. Similar to the GAMLSS method, we can estimate a symmetric subgroup of regression quantiles (percentile curves).

It is a robust estimation of the parameters of a linear regression model. The estimation procedure is similar to an ordinary least squares (OLS) estimate for linear regression, where you minimise the sum of squared residuals to find the best-fitting line for the mean. We assumed that the relationship between the p quantile of Y and covariate t is given by

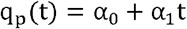

Given the dataset 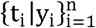, We estimated the parameters α by minimizing

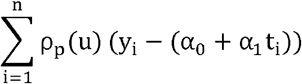

Where ρ_p_ (u) = |u| is the check function (or loss function). Here, we minimised the sum of weighted absolute residuals to find the best-fitting line for a specified quantile. The check function can be rewritten as

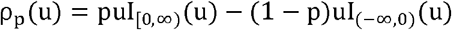

Where I_A_ (u) is the indicator function of set A written as

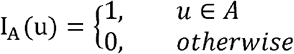

However, there is no explicit solution for the regression coefficients since the check function is not differentiable at the origin. Portnoy and Koenker (1997) discussed and implemented the modified version of the Barrodale and Roberts algorithm to solve this computation problem ^30^. It is the default method in the R package quantreg to estimate the parametric regression quantiles.

### Nonparametric method without distributional assumption: HRY

The HRY method is a two-step modelling technique to calculate centiles without distributional assumptions.

The first step is a sliding window approach.

- Sorting the data by time: First, the observed measurements Y (here stroke severity) are sorted in ascending order of time t (time from symptom onset to assessment).
- Selecting a subset to run a local regression: A subset of k measurements of Y is selected, where k is a fraction (typically 5-10%) of the total number of observations. This subset is used to perform a regression, here of admission stroke severity on time.
- Estimating residuals: The selected k measurements are regressed on *t* by a simple linear regression with OLS estimation, and the residuals (the differences between observed and predicted values from the regression) are calculated.
- Calculating centiles: We sort the residuals, and determine the centiles (e.g., 10th, 50th, 90th) based on this distribution. Then we plot these centiles against the median time of the first k observations. The residuals effectively “centre” the data around zero (the predicted admission stroke severity), so when we calculate the centiles of these residuals, we measure the spread of stroke severity scale around this local average. By adding these centiles back to the predicted admission stroke severity from the regression, we can estimate the actual admission stroke severity centiles for that time of assessment.
- Plotting the centiles: When we plot the centiles against the median time of the window, we see how these stroke severity centiles vary with time. Since each window is small and overlaps with others, these plots will form a successive but rough curve when combined.
- Sliding window: The procedure is repeated by shifting the subset by one position, i.e., using points 2 to k+ 1, 3 to k+ 2, and so on, until the entire time range has been covered. This sliding window process ensures that all data points are used in the centile estimation.
- Edge effects: Because this method involves averaging over a window, approximately 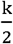 data points at each end of the age range need to be omitted from the final centile plots, as these points do not have a full window of k observations to average over.

This ensures that the centile estimates are based on well-centred windows, where the local regression is more representative of the true relationship at that point.

The second step is to use polynomials with a degree of p to smooth the centiles.

- Smoothing each centile curve: Fit a smooth curve to each centile (percentile) of the data. The centile curves are assumed to be described by a polynomial function of time t. And the polynomial function of degree p is given by

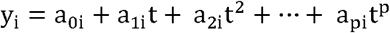

Where y_i_ is the smoothed value of the i th centile, and a_ji_ are the coefficients of the polynomial, which are specific to each centile i. The degree *p* determines the flexibility of the curve. *p*= 2 is typically sufficient for short spans of *t*, as it captures quadratic trends. For stroke severity, where changes over time might exhibit non-linear dynamics (e.g., due to evolving symptom severity), experimenting with higher p values may be necessary.

- Smoothing across centiles: Ensure that the intervals between centiles at a fixed time also behave smoothly. We write the coefficients of the polynomial a_pi_ to be the polynomial functions of z_i_. The z_i_ represents the normal equivalent deviate (NED) of the i th centile. For example, the 50th centile (median) has a P_i_ = 0.5, and its corresponding z_i_ = 0. For lower and higher centiles, z_i_ will be negative and positive, respectively. And the polynomial function of degree *q* is given by

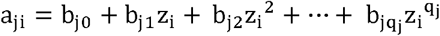

Here, q_j_ is the degree of the polynomial for each coefficient a_ji_. Higher q_j_ allows more flexibility. For example, q_0_ = q_1_ =1 with the other q values equal to zero assumes the intervals between centiles are Gaussian (uniform spacing). q_0_ =2 accounts for skewed distributions. q_0_ =3 accommodates distributions with long tails.

- Combining the two smoothing process: By substituting the polynomial form of a_ji_ into the initial polynomial equation for y_i_, we get the final model

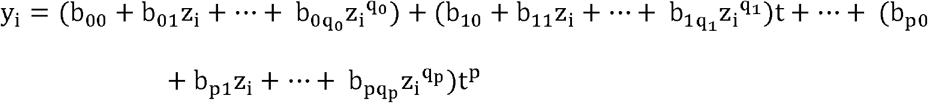
- Fitting the model: The final model, which combines the smoothness across times (within centiles) and across centiles (at any given time), can be fitted using the least squares method. This statistical method minimises the sum of the squares of the differences between the observed and predicted values.
- Selecting parameters *k, p* and q_j_ in the model: We evaluated *k* to be 10% of the total number of observations as a balance of overfitting and flexiblity. We considered *p*= 2 (quadratic) as this should adequately capture changes in severity over time. We used q_0_ =2 to handle skewness in centile intervals, and set q_1_ = 1, q_2_ =0 for simplicity in higher-order terms. See details of paremeter selection in Appendix D and E. The final model can be written as

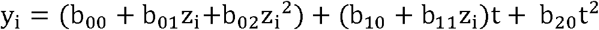

We still want to estimate a symmetric subgroup of percentile curves, such as the 10th, 20th, 30th, 40th, 50th, 60th, 70th, 80th, and 90th percentiles. The P_i_, NED and final model equation for each centile is given in Table 2.

**Table 2.** Summary of model details for the smoothed centiles.

| Centile | $P_i$ | NED | Model equations |
| --- | --- | --- | --- |
| **10th** | 0.1 | -1.28 | $y_{10} = (b_{00} - 1.28b_{01} + 1.64b_{02}) + (b_{10} - 1.28b_{11})t + b_{20}t^2$ |
| **20th** | 0.2 | -0.84 | $y_{20} = (b_{00} - 0.84b_{01} + 0.71b_{02}) + (b_{10} - 0.84b_{11})t + b_{20}t^2$ |
| **30th** | 0.3 | -0.52 | $y_{30} = (b_{00} - 0.52b_{01} + 0.27b_{02}) + (b_{10} - 0.52b_{11})t + b_{20}t^2$ |
| **40th** | 0.4 | -0.25 | $y_{40} = (b_{00} - 0.25b_{01} + 0.06b_{02}) + (b_{10} - 0.25b_{11})t + b_{20}t^2$ |
| **50th** | 0.5 | 0.00 | $y_{50} = b_{00} + b_{10}t + b_{20}t^2$ |
| **60th** | 0.6 | 0.25 | $y_{60} = (b_{00} + 0.25b_{01} + 0.06b_{02}) + (b_{10} + 0.25b_{11})t + b_{20}t^2$ |
| **70th** | 0.7 | 0.52 | $y_{70} = (b_{00} + 0.52b_{01} + 0.27b_{02}) + (b_{10} + 0.52b_{11})t + b_{20}t^2$ |
| **80th** | 0.8 | 0.84 | $y_{80} = (b_{00} + 0.84b_{01} + 0.71b_{02}) + (b_{10} + 0.84b_{11})t + b_{20}t^2$ |
| **90th** | 0.9 | 1.28 | $y_{90} = (b_{00} + 1.28b_{01} + 1.64b_{02}) + (b_{10} + 1.28b_{11})t + b_{20}t^2$ |

### Dataset and sample

The dataset was constructed using the admission stroke severity values at a single stroke unit, in the Department of Neurology, Haukeland University Hospital, between February 2006 and February 2021. The patient data was pseudonymised and obtained by Haukeland University Hospital with approved informed consent. Participants data were retrospectively collected from their electronic patient records in the hospital-registered database.

Stroke severity was assessed by the National Institutes of Health Stroke Scale (NIHSS) at admission. Symptom onset is when the patient or a reliable observer can identify the moment stroke symptoms began. When the time of symptom onset is unknown, the time the patient was last seen or felt well (before symptom onset) is used as a proxy ^31^. Assessment time was recorded as the moment when the clinical team conducted the initial NIHSS evaluation. The interval between symptom onset and assessment was calculated as the differential between these two time points.

We excluded patients without recorded NIHSS; with undocumented symptom onset time, transient ischaemic attack diagnosis, unspecified stroke types, or symptom onset beyond 12 hours. We selected a 12-hour time window because we want to take into account most of the patients who had their admission NIHSS assessed within the hyperacute stage of stroke. The relationship between the time from symptom onset to assessment and admission stroke severity may differ depending on the stage of post-stroke recovery, i.e., hyper-acute, acute, and rehabilitation phases ^32^. The majority of stroke patients, however, present during the hyper-acute and acute stages when symptoms are rapidly evolving and treatment decisions are most time-sensitive.

After the data cleaning and patient screening process in Appendix A’s flowchart (Figure 5), we applied and compared the above three methods separately for patients with ICH and IS.

## Results

### Study sample characteristics

We summarised the patient characteristics of the study sample in Table 3. A total of 2040 IS patients were included with a median (IQR) age of 74 (62 to 82) y; 43% were male. The median (IQR) admission severity scale was 3 (1 to 8), and the median (IQR) time to assessment was) 2.3 (1.3 to 4.3) hrs, respectively. A total of 272 patients with ICH were included. There were fewer male (41%) and older age (median (IQR) 75 (65 to 81)) patients with more severe strokes (median (IQR) 11 (4 to 18)) on admission. The patients with ICH were admitted to the hospital and undertook the admission stroke severity assessment more quickly (median (IQR) 1.9 (1 to 3.2) hrs) than ischaemic stroke patients. The differences in patient characteristics, especially admission severity and time to assessment, highlight the need to model the centiles separately for different stroke types.

**Table 3.** Patient characteristics of the study sample.

| **Characteristics** | **IS**<br>**n=2044** | **ICH**<br>**n=273** |
| --- | --- | --- |
| **Demographic** |  |  |
| Age, median (IQR) y | 74 (62 to 82) | 75 (65 to 81) |
| Sex, (Male=1) % | 43 | 41 |
| Time from symptom onset to admission NIHSS assessment, median (IQR) hrs | 2.3 (1.3 to 4.3) | 1.9 (1 to 3.2) |
| Admission NIHSS, median (IQR) | 3 (1 to 8) | 11 (4 to 18) |
| **Medical history** |  |  |
| Pre-stroke mRS, Median (IQR) | 0 (0 to 1) | 0 (0 to 1) |
| Cerebral infarction % | 12 | 9 |
| Subarachnoid haemorrhage % | 1 | 4 |
| Heart attack % | 15 | 11 |
| Angina % | 14 | 11 |
| Peripheral vascular disease % | 7 | 6 |
| Hypertension % | 53 | 52 |
| Diabetes % | 14 | 14 |
| Paroxysmal atrial fibrillation % | 11 | 11 |
| Chronic atrial fibrillation % | 9 | 10 |

### Comparisons of estimated centile curves

#### Admission stroke severity centile curves for ischaemic stroke

Generally, the estimated centile curves from different methods agree regarding shape and magnitude for IS patients in Figure 2. However, there were marked differences.

**Figure 2.**
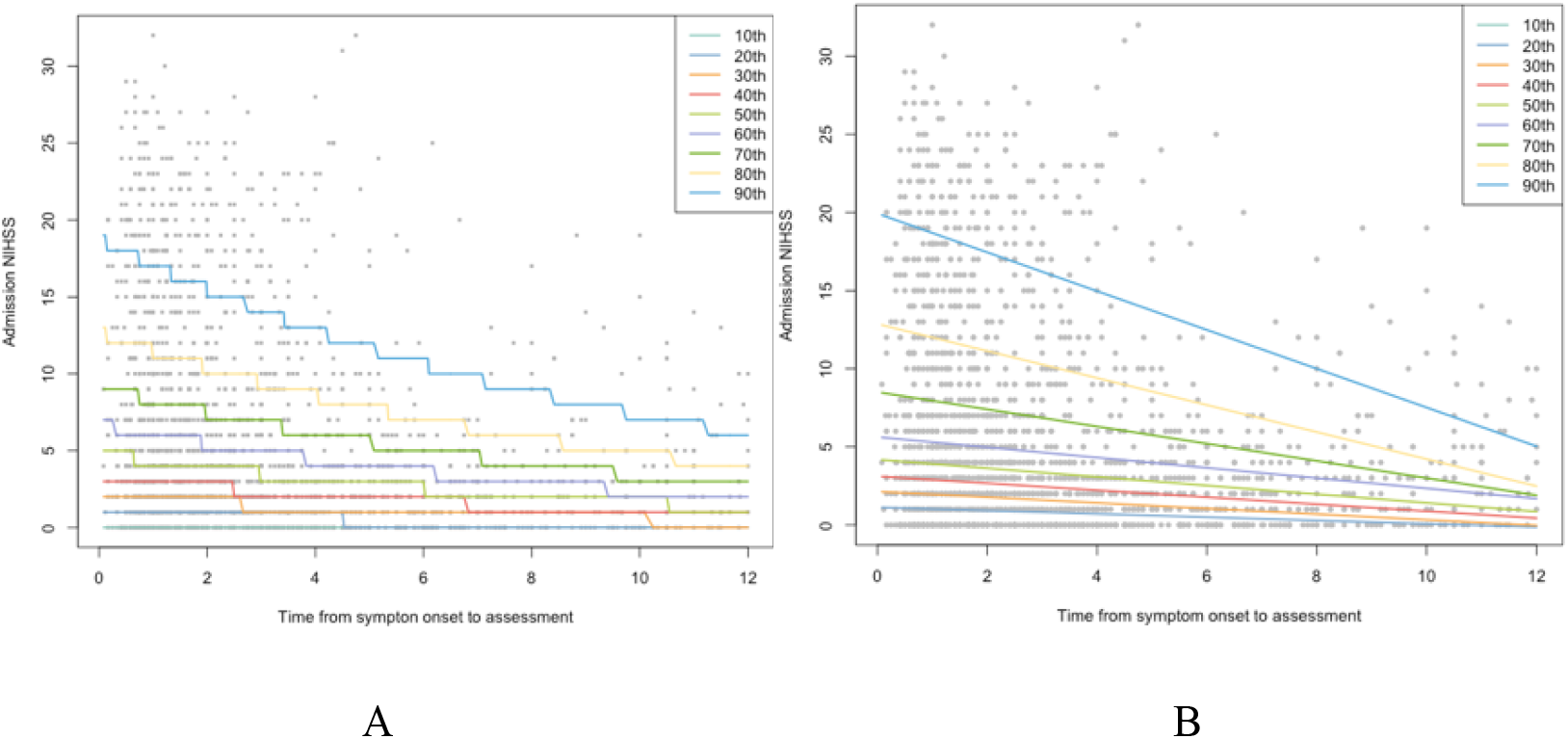
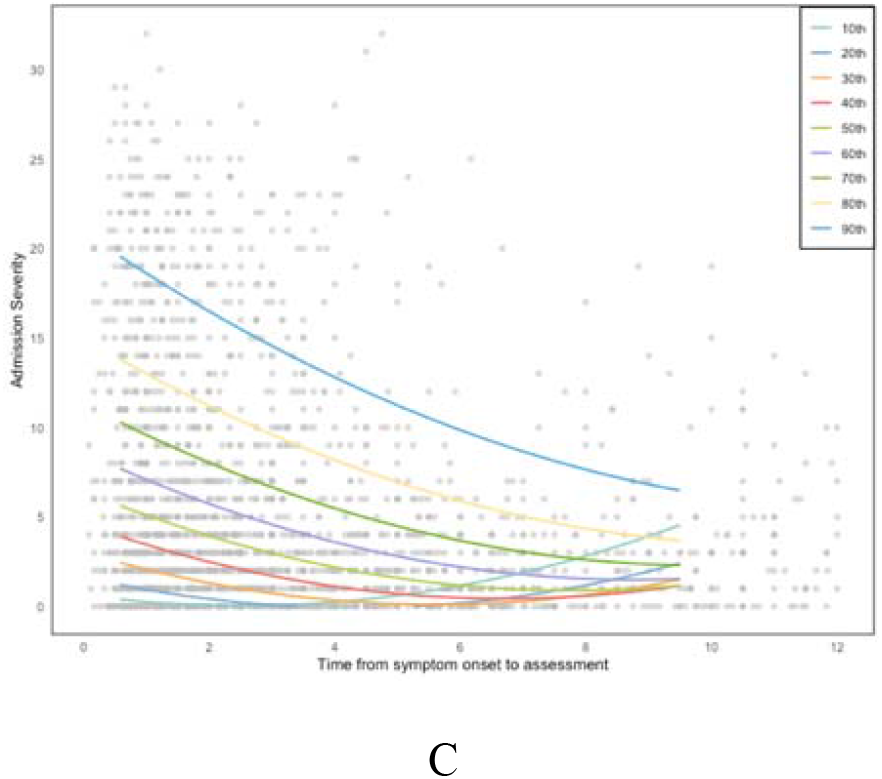
Centile curves modelled for IS. (A) GAMLSS fitted with a negative binomial distribution; (B) QR; (C) HRY

The centiles estimated by GAMLSS may better reflect the actual stroke severity scale as non-negative integer values. The shape of the centile curves was stepped. GAMLSS model fitted with negative binomial distribution and linear additive terms achieved the best performance. The complete model diagnosis, selection and parameter estimation process is summarised in Appendix B and C.

The centile curves estimated by QR are smoothed as the admission stroke severity scale was treated as a continuous response variable in the models. We assumed a linear relationship between admission stroke severity and time from symptom onset to assessment, so the centile curves estimated by QR are straight lines. The overlap in quantile regression (QR) centile curves occurs because each quantile was estimated independently without constraints, ensuring higher quantiles (e.g., 90th) always lie above lower ones (e.g., 10th), especially in areas with sparse data. This issue was exacerbated by small sample sizes and high variability at later times post-symptom onset, where fewer data points can support the estimates. While larger sample sizes would reduce overlap by stabilising quantile estimates, minor inconsistencies may persist due to the lack of monotonicity assumption. We also cannot estimate some lower centiles, such as the 10th centile by QR, as we had zero values in admission stroke severity. This was because, for the 10th percentile (or 0.1 quantile), the regression tried to find the line where 10 per cent of the observed data points fall below the predicted values and 90 per cent fall above. If all the values at the 10th percentile are zero, there’s no variation in the data. The design matrix becomes singular. So, we cannot estimate the parameters as this prevents the algorithm from finding a unique solution.

In the HRY method, the centile curves were smoothed by polynomial functions of degree two, so they are nonlinear. The “uptick” in the estimated centile curves was observed at later time points. This even caused the centile curve to cross in the lower centiles, such as the 10th, 20th, 30th, and 40^th^. The excessive zero values were likely drivers of the observed “uptick” in the lower centiles for IS patients despite the centiles being smoothed jointly to avoid crossing. In the first step of the HRY method, local regression was used to estimate residuals and centiles. When the data contain excessive zero values, particularly in the lower centiles, the regression was heavily influenced by the non-zero values. If the non-zero values dominate the local regression fit, they can distort the shape of the centile curves, especially in regions where zero values are interspersed with sparse high values. In the middle time intervals, the regression, dominated by a mix of zero and moderate values, produces negative estimated intercepts. In the later intervals, the appearance of more high values reversed this trend, leading to positive estimated intercepts. This distortion and excessive zero values drived the “uptick” in the lower centiles at later time points. We also cannot identify the stroke severity centiles for patients who had NIHSS assessed early and late, in our case, about the first 0 to 0.6 hours and the last 9.5 to 12 hours for IS patients. As mentioned in the method section, the first step of HRY method omitted 5 per cent data points to run the local regressions and it cannot provide estimation at the two ends. This ensured that the centile estimates were based on well-centred windows, where the local regression was more representative of the true relationship at that point.

#### Admission stroke severity centile curves for ICH

The estimated centile curves from different methods varied in Figure 3 for ICH patients. The centiles at the ends were not precisely estimated due to the small sample size for ICH patients. For the GAMLSS method, the ICH centile curves maintained an overall shape that is comparable to those for IS, reflecting a consistent trend in the relationship between stroke severity and time from symptom onset. However, the curves for ICH demonstrated a larger variation across the centiles, aligning with the clinical understanding that ICH tends to present with greater severity compared to IS. The larger variation was because we assume one frequency distribution and estimate the distribution parameters (moments), and the estimation procedure borrows strength across the time range.

**Figure 3.**
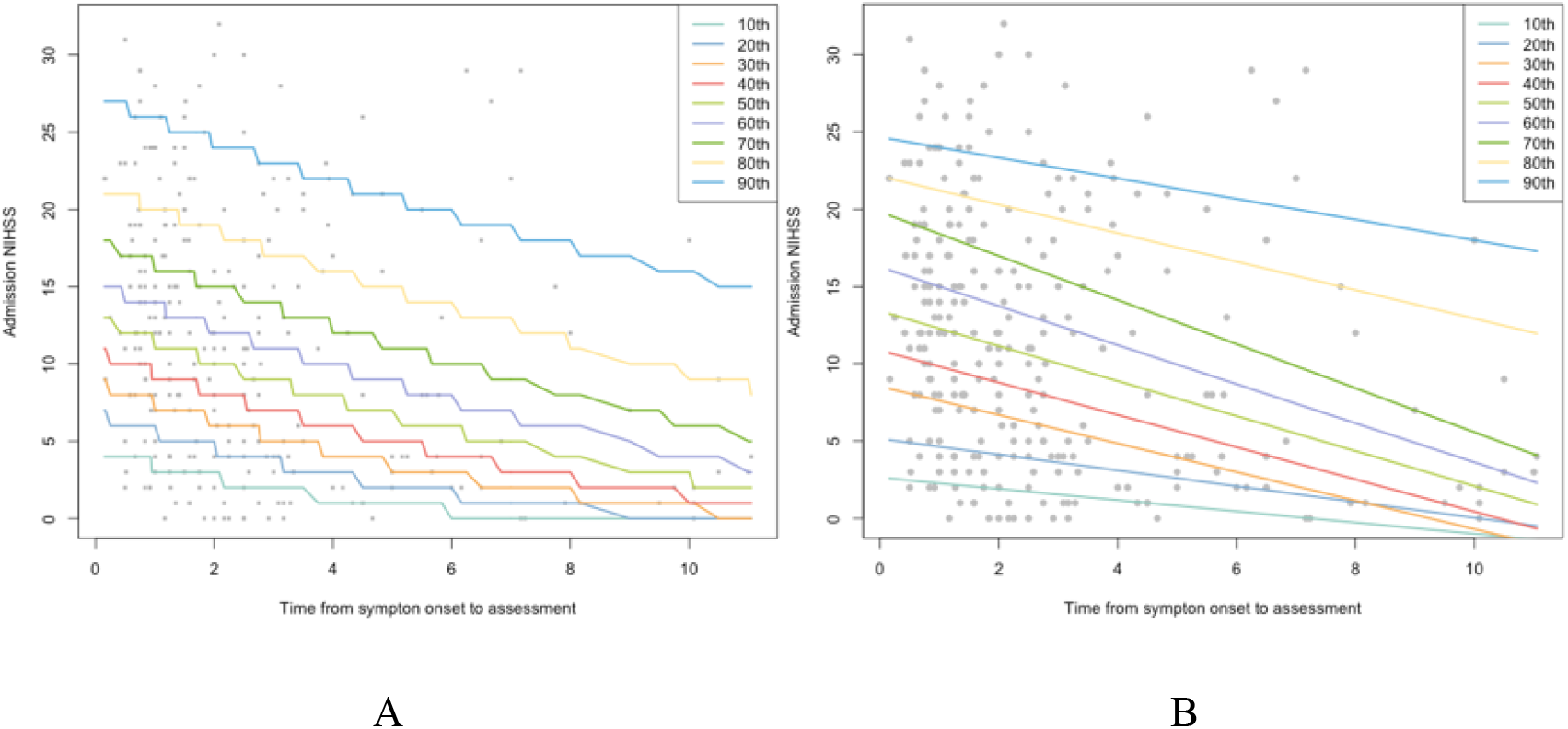
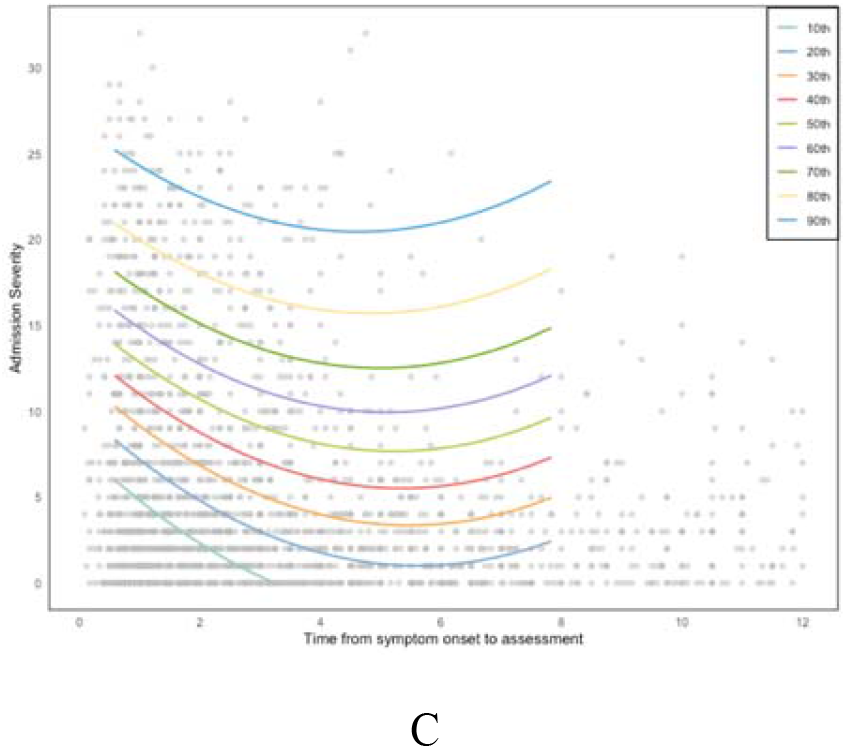
Centile curves modelled for ICH. (A) GAMLSS fitted with a negative binomial distribution; (B) QR; (C) HRY

The centile curves estimated by QR are crossed near the extremes because those curves were supported by even fewer observations compared with the IS cohort. However, we can estimate the 10^th^ centile curve for ICH patients as there were fewer zero values of NIHSS compared with IS patients, enabling the parameter estimation.

The shape of centile curves estimated by HRY methods also showed an ‘uptick’ but for all the curves. The local regressions at the ends are very underpowered due to the small sample size and are less representative of the true relationship. Similar to IS patients, we cannot identify the stroke severity centiles for patients with NIHSS assessed in the first 0 to 0.6 hours and the last 8 to 12 hours.

### Clinical interpretations

#### Stroke severity is not a static baseline factor

Instead of using a single assessment to determine patient stroke severity on admission, we adjusted for the time from symptom onset to assessment after modelling the centiles to represent the temporal dynamics of stroke severity. We are interested in the position of a patient’s stroke severity relative to other patients’ stroke severity who were admitted to the hospital at around the same time after symptom onset. In this case, the same score is interpreted in terms of severity level relative to patients admitted to the hospital at the same time after symptom onset.

Take the centile curves estimated by GAMLSS method as an example for interpretation, as shown in Figure 4 A, an IS patient with admission NIHSS scored a six is on the 60^th^ percentile at one hour of admission after symptom onset. This means that 40% of the patients admitted to the hospital one hour after symptom onset have a higher stroke severity scale. A patient with the same score of six is on the 70^th^ percentile four hours after symptom onset and the 80^th^ percentile eight hours after symptom onset. Therefore, the same stroke severity score for a patient is in a higher position (more severe) relative to other patients’ symptom severity for later admission times.

**Figure 4.**
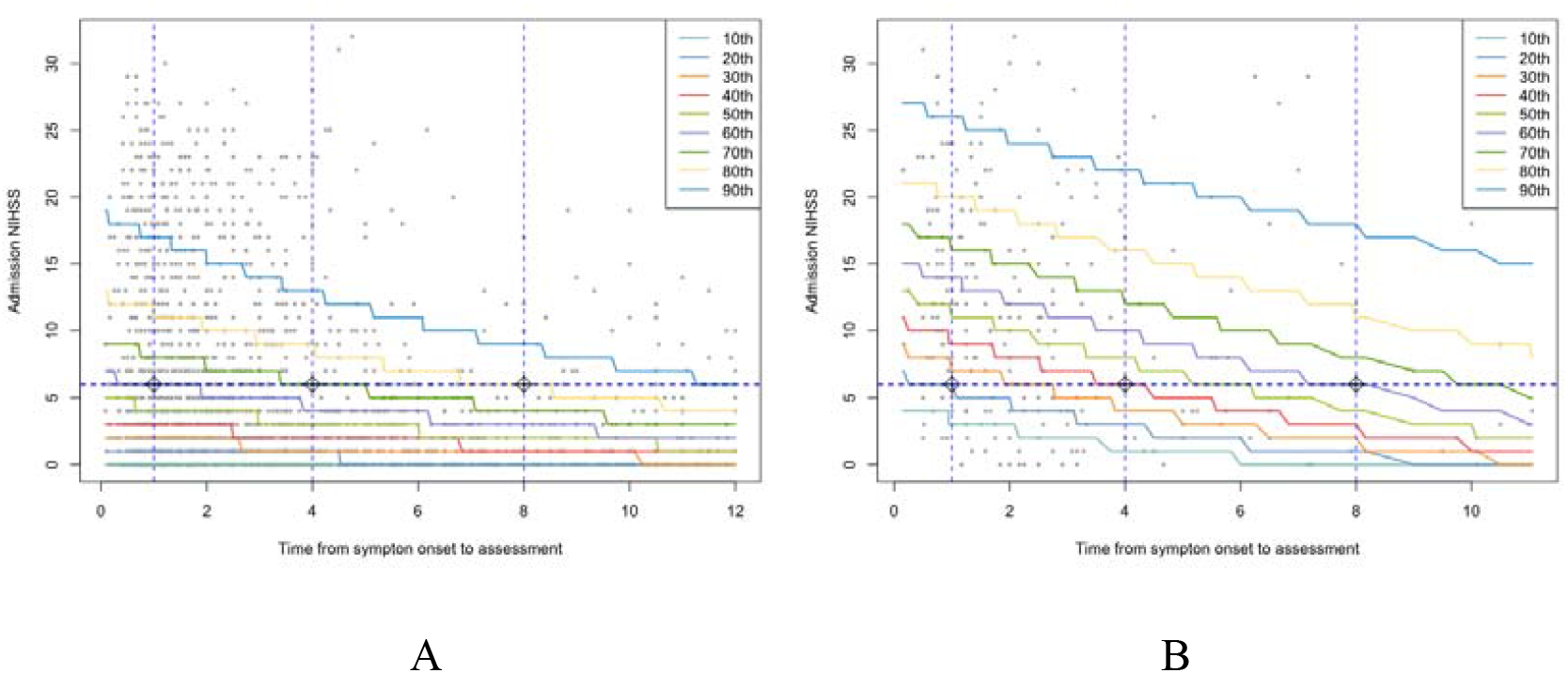
Interpreting stroke severity based on centile curves modelled by GAMLSS fitted with a negative binomial distribution for IS (A) and ICH (B)

**Figure 5.**
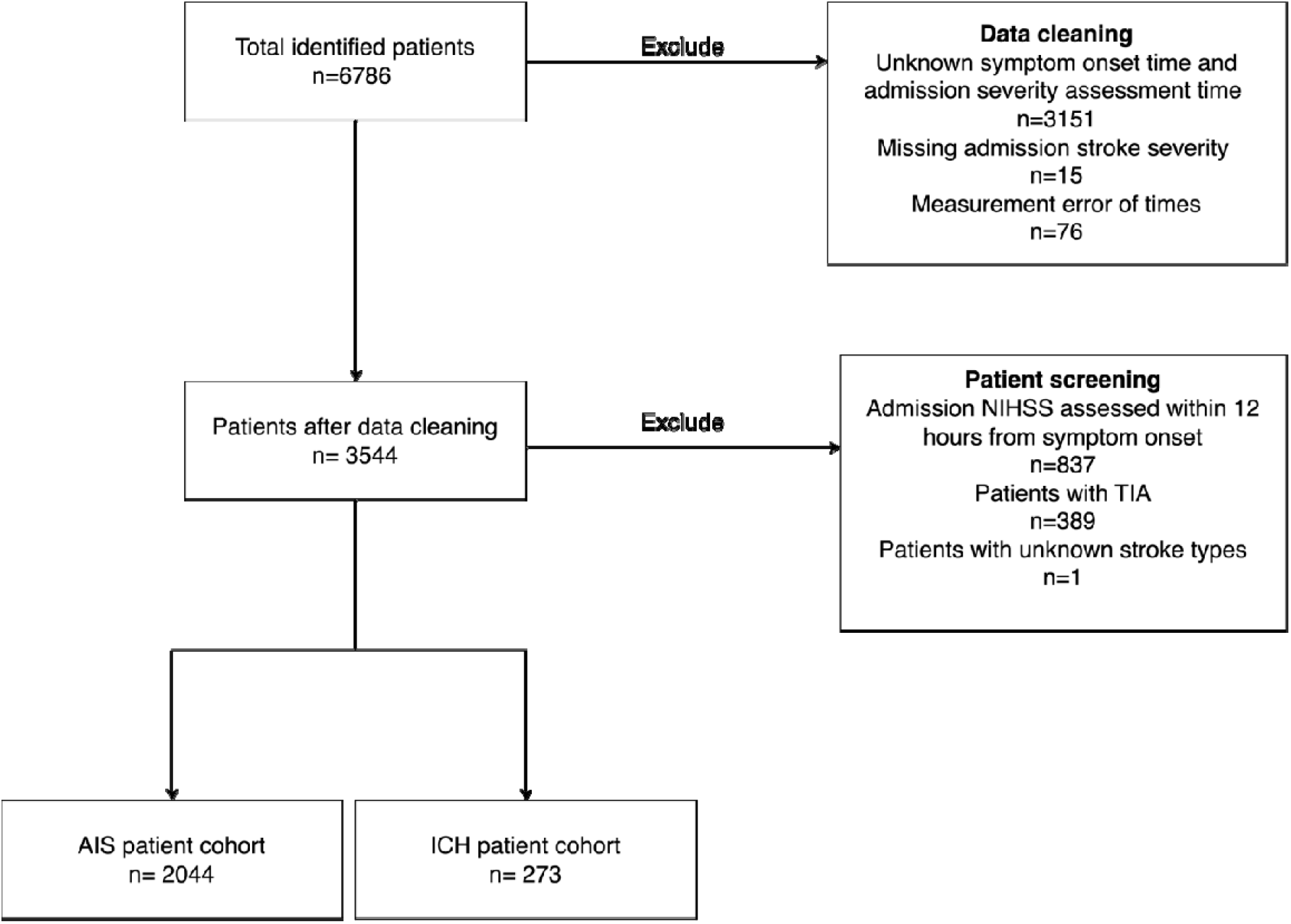
Flowchart of data cleaning and patient screening

### Centile curves for different stroke types

The interpretation of stroke severity centiles is also different between ischaemic stroke and ICH. From Figure 4 B, patients with ICH were more severe on admission compared with patients with IS. Again, a patient with an NIHSS severity score of six on admission is only on the 20^th^ percentile at one hour of admission after symptom onset, 40^th^ percentile at four hours of admission, and 60^th^ percentile at eight hours of admission. Stroke severity for ICH also changed more rapidly compared with ischaemic stroke. This can be observed from the estimated centile curves as they step more steeply. For example, the centile changes at each hour of delayed admission in a haemorrhagic stroke patient with the same severity score of six on admission, as presented in Table 4.

**Table 4.** The percentile of NIHSS=6 given specific assessment time for IS and ICH patients.

| Assessment time (hrs) | IS | ICH |
| --- | --- | --- |
| **1** | 60th | 20th |
| **4** | 70th | 40th |
| **8** | 80th | 60th |

### Clinical implications

#### Classification of stroke severity scale

The stroke severity scale can be classified when presenting summary statistics of baseline patient characteristics or comparing patient outcomes. The current practice of defining categories is only based on a single stroke severity score measured at admission ^33, 34, 35^. The common classifications are (1) Three groups: mild (0–6), moderate (7–15), or severe (>15) (2)

Four groups: mild (<5), moderate (5-15), severe (15-20) or very severe (>20), and (3) Five groups: no stroke symptoms (0), minor stroke (1–4), moderate stroke (5–15), moderate to severe stroke (16–20), and severe stroke (21–42). After modelling the centiles, we can use a continuous, percentile-based scale rather than fixed categorical definitions so that severity can be relative to the time of assessment, rather than a strict absolute severity measure. For example: mild (0-25^th^ percentile), mild to moderate (25^th^ -50^th^ percentile), moderate to severe (50^th^ to 75^th^ percentile) and Severe (75^th^ to 100^th^ percentile). This flexible classification allows you to shift severity categories dynamically based on the population distribution, making it more responsive to trends in the data (e.g., more severe strokes in a certain cohort).

### Patient selection in RCTs

Current standard practice within RCTs associated with stroke is to select eligible patients based on admission stroke severity; for example, the Interventional Management of Stroke (IMS) Phase III trial used admission NIHSS>10 as one of the inclusion criteria ^36^. After modelling centiles, we may reconsider whether we need to select patients based on their severity level relative to others at a given time, as this better reflects the patient’s condition taking into account the underlying admission process. For example, we may select patients with admission stroke severity over 80^th^ centiles. Also, given the dynamic nature of stroke severity, we may consider a second measure before randomisation to capture any changes in symptom severity, although enrolment into such trials and initiation of treatment is generally time-limited.

### Patient monitoring and treatment

The stroke severity centiles might be useful in informing clinical treatment decisions. Admission NIHSS is currently used to select patients eligible for specific treatments, particularly in the ‘hyper-acute’ period of the first 0 to 24 hours ^32^. For example, NIHSS ≥ 6 is used to identify eligible patients for further assessment to undertake mechanical thrombectomy within 24 hours ^37^. This period is when centiles are steepest, and it may be that more appropriate targeting of those with the potential to benefit from such treatments is possible by using severity centiles rather than raw severity scores.

### Stroke prognostic models

There is a more direct implication in informing stroke prognostication. The current research practice is to use admission stroke severity as a fixed baseline covariate in prognostic models ^11^; a more severe stroke on admission is usually associated with worse outcomes ^38^. However, incorporating severity centiles in place of raw scores could improve the efficiency of prognostic models, which may lead to better patient outcome predictions. This can be further applied to increase the power of an RCT to adjust for prognostic variables when conducting the statistical analysis for trials ^39^.

## Discussion

In this paper, we constructed the admission stroke severity centiles using GAMLSS, QR and HRY methods. Centile modelling is an approach to adjust the time from symptom onset to assessment as a covariate. We expressed the measurement of stroke severity for individual patients at admission as estimated centiles rather than its original scales. In the context of centiles, we interpreted admission stroke severity with additional information on time. Using this approach, patients who score the same on admission are considered to have different stroke severity depending on the time to assessment since symptom onset. Admission stroke severity centiles have various potential clinical implications, such as optimising stroke severity categories, enhancing clinical trial design, informing clinical stroke care and improving patient prognostication.

We also showed that it is possible to model centiles for scale measurements with typically skewed distribution. They are not approximately normally distributed like many continuous anthropometric measurements ^40^. We selected and applied three methods to estimate centiles; all three methods have advantages and disadvantages. The GAMLSS method provides a comprehensive modelling framework to fit different distributional assumptions, evaluate model fit and select optimal models with an explicit estimated density function to allow the direct centile computation without crossing. It is less sensitive to outliers; extreme centiles such as the 10th and 90th are more stable. However, finding a distributional assumption to fit the stroke severity scale is challenging. Considering the stroke severity scale as a count variable is reasonable but not ideal ^41^.

Quantile regression has more limitations than the GALMSS method. The quantile curves near the extremes are more erratic and may cross as they are estimated separately. The fitted quantile regression model does not have a (likelihood-based) measure of fit to construct the likelihood ratio test, AIC, or BIC. This creates difficulties in model selection.

HRY allows considerable flexibility in modelling the centiles, and compared with other computationally intensive nonparametric methods, it is easy to understand and implement. However, it cannot estimate the centiles from two ends. Stroke symptom severity can change dramatically in patients admitted to the hospital early, so the inability to estimate centiles at the left end is a major disadvantage. There are also no R packages available for the HRY method. It is also difficult to define the residuals of a fitted quantile regression model for model diagnosis. The model selection is only based on informal experimentation of different sets of parameters of *k, p* and q_j_. The choice of parameters in the HRY method is intuitive.

Larger values of *k* create smoother centile curves but may fail to capture localised variations in data. Smaller values of *k* capture local changes but can lead to noisy or overfitted centile curves. We also evaluated *k* = 0.05. We also tried *p*= 1 (linear) and *p*= 3 (cubic). We did not consider *p*> 3 to avoid overfitting. Full results are displayed and discussed in Appendix D and E.

Overall, we prefer the GAMLSS method and recommend using it to construct admission stroke severity centiles. GAMLSS accommodates the discrete and skewed nature of stroke severity data, ensures a robust estimate of centile curves, and avoids the issues of overlapping or crossing centiles observed with QR and HRY.

GAMLSS is not an ideal solution; other available methods have not been applied in this study ^15,16^. This paper aimed to demonstrate the feasibility and practicability of modelling stroke severity centiles. Future research may focus on reviewing and seeking the best method. Other methods were rejected because they rely on strict normality assumptions, require data transformation to achieve normality, lack of solid model fit evaluation framework, or are rarely implemented and difficult to interpret outside the statistical or methodological fields.

The estimated centile curves of admission stroke severity in this study cannot currently be used as a reference chart. It instead can only be considered an internal adjustment in this study sample to interpret stroke severity in the context of when patients come to the hospital. These centiles are not estimated by a large sample to represent the characteristics of the population regarding admission stroke severity in a specific area, country or region. The recommended sample size is 7500 - 25000 to construct a birthweight reference centile chart to reach the prespecified minimum precision ^42^. However, further investigation is needed to see if this is the case for constructing reference centiles of the admission stroke severity.

Our participants were from a single site, and there were too few on whom to base reference centiles for global use. However, we have shown the feasibility of the methods and discussed the potential uses of such reference centiles. Further work should also consider how to incorporate patients with unknown onset time.

## Data Availability

All data produced in the present study are available upon reasonable request to the authors

## Appendices

### Appendix A Data cleaning and patient screening

We had a total of 6786 patients in the original dataset. In the initial data cleaning process, we excluded 3151 patients with missing time of symptom onset and stroke severity assessment and 76 patients with mistakes in time recording. We also excluded 15 patients who had their stroke severity missing at admission. Based on the eligibility criteria, we further excluded 837 patients with admission stroke severity after 12 hours from symptom onset, 389 patients with TIA and one patient with unknown stroke subtypes. After data cleaning and screening, we had 2044 IS patients and 273 ICH patients included separately to construct the centiles.

### Appendix B GAMLSS Model fitting, diagnosis and selection

There is only a small improvement in model fit by fitting three parameters’ distributions in the GAMLSS model. For IS patients, the Rigby and Stasinopoulos algorithm for parameter estimate did not converge when fitting beta-binomial distribution. We used the Cole and Green algorithm instead ^4^. When fitting the Delaporte distribution, both the Rigby and Stasinopoulos algorithm and the Cole and Green algorithm failed to converge. We used a mixed algorithm by using the RS algorithm twice before switching to the Cole and Green algorithm for up to ten extra iterations. For haemorrhagic stroke patients, similar issues also appeared. Therefore, we used a mixed algorithm when fitting Delaporte and beta-binomial distributions.

We used the worm plot for model diagnosis and applied the wp() function from the GAMLSS package. The worm plots identify the intervals of the explanatory variable within which the model does not adequately fit the data. The data points in each plot form a worm-like string. The shape of the worm indicates how the data differ from the assumed underlying distribution. A flatworm indicates that the data follow the assumed distribution in that time interval ^43^.

The selection of GAMLSS models is based on how well we fit the data, which is mainly judged by generalised Akaike information criteria (GAIC). GAIC is defined by the fitted global deviance, the effective degree of freedom used for the fitted model, and a penalty for each degree of freedom used. There are two special cases in GAIC: Akaike information criteria (AIC) and Bayesian information criteria (BIC) ^44^. We used BIC to select the nested models with and without additive terms under the same distributional assumption and the unnested models under different distributional assumptions.

From the worm plots in Figure 6 for IS patients, the model fit of centile estimation did not change much according to different distributional assumptions. There are approximately flat worms across 12-time groups, especially for the first four hours. The additive terms of cubic splines and fractional polynomials show better model fit than linear assumption, especially after the first 4 hours. However, worm plots of the centile model for ICH patients in Figure 7 showed an S-curve, indicating a lack of fit for the chosen distributions on the data in modelling the changes of skewness and kurtosis.

**Figure 6.**
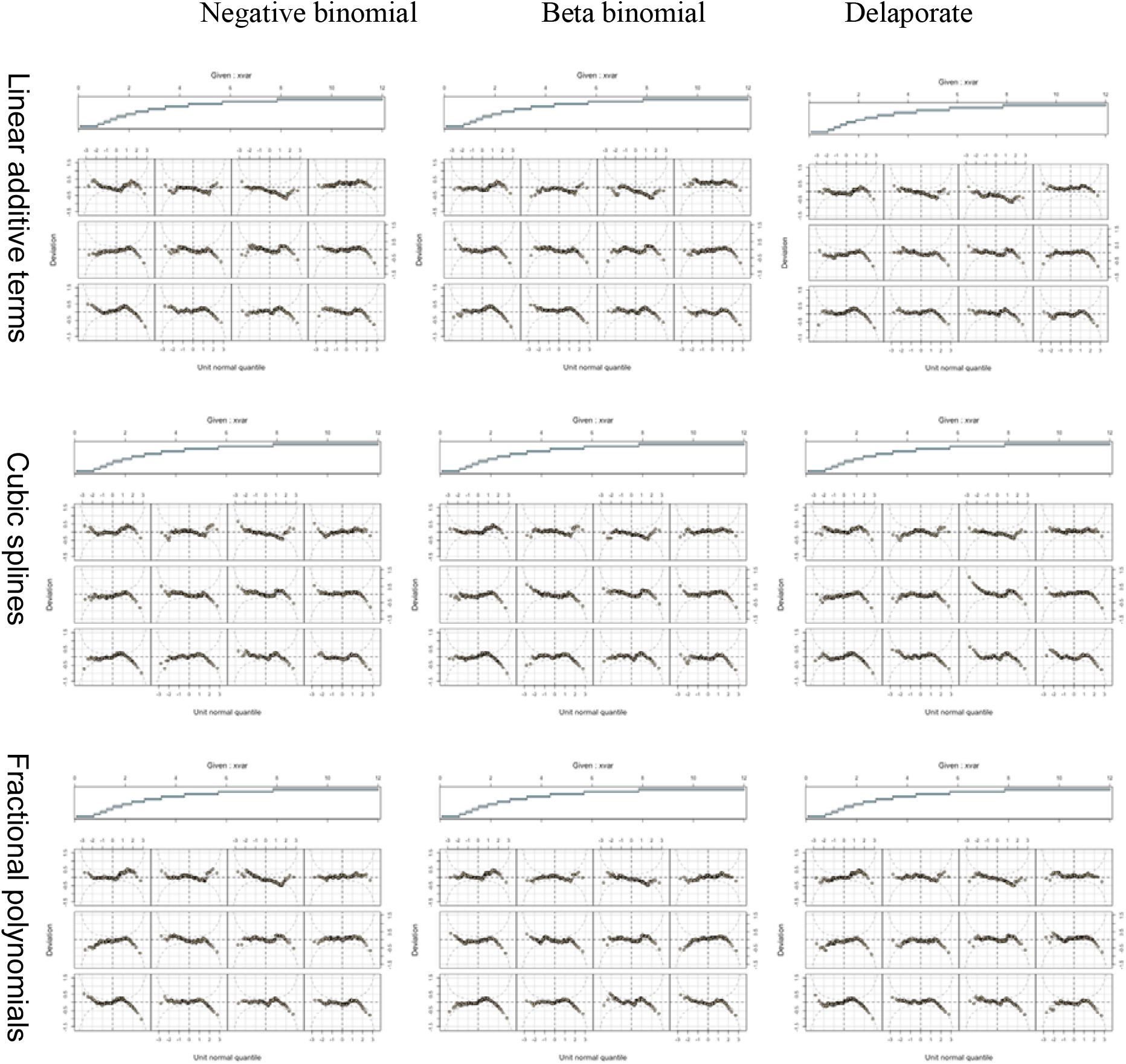
GAMLSS model diagnosis based on worm plot: admission stroke severity centile for IS patients

**Figure 7.**
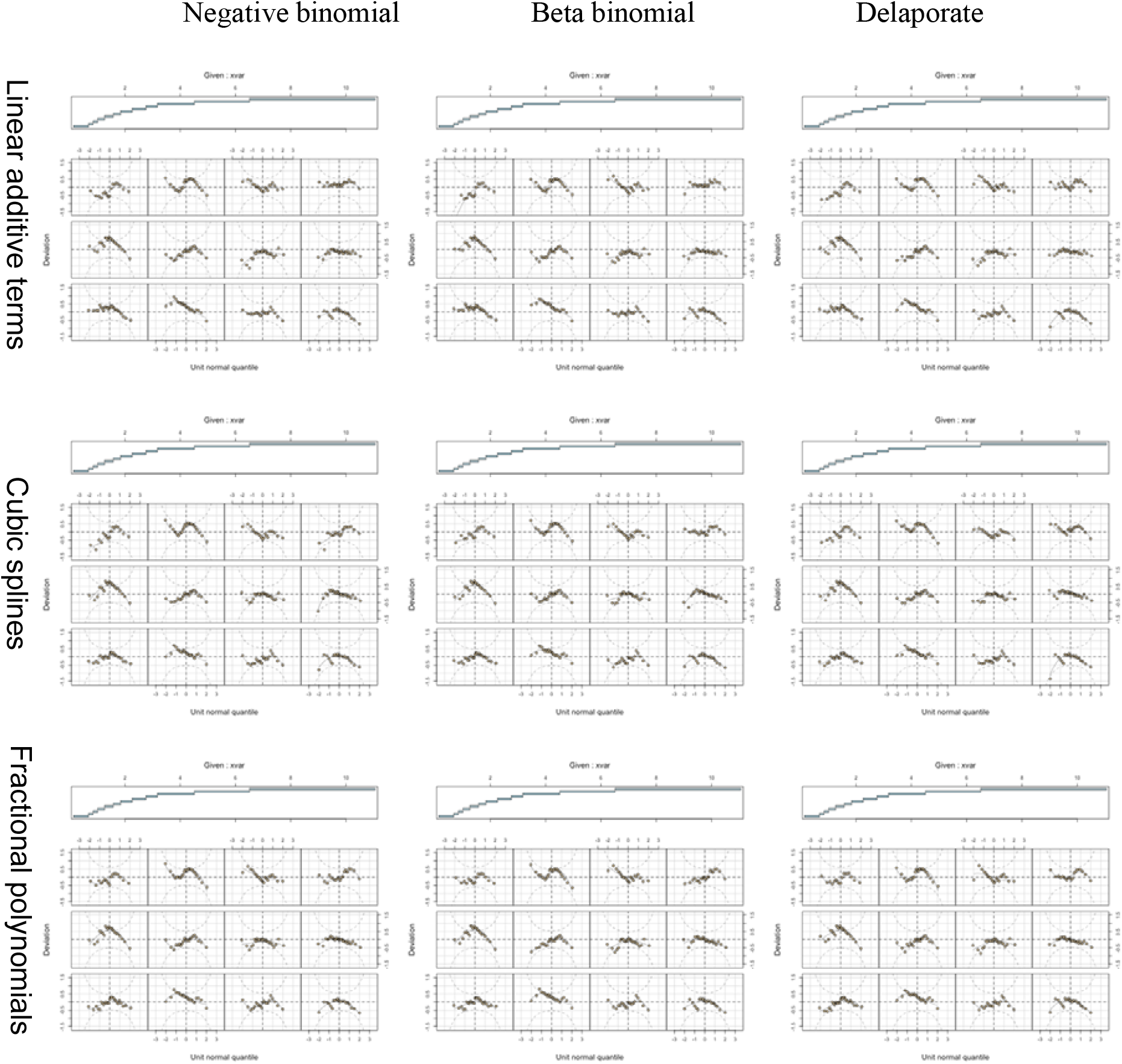
GAMLSS model diagnosis based on worm plot: admission stroke severity centile for ICH patients

For selecting the final centile model in Table 5 and Table 6, the negative binomial model with linear additive terms appears to be the best based on BIC for both IS and ICH patients. We did not select models assuming nonlinear additive terms, although they achieved a better fit as indicated by the worm plot. They captured the noise instead of the characteristics of the dataset. We can observe the fact as well from the estimated parameters in Appendix C below.

**Table 5.** GAMLSS Model selection based on BIC: admission stroke severity centile for IS patients.

| **Distributions**<br>**Additive terms** | **Negative binomial** | **Beta binomial** | **Delaporte** |
| --- | --- | --- | --- |
| **Linear** | **11454** | 11470 | 11465 |
| **Cubic splines** | 11464 | 11504 | 11494 |
| **Fractional polynomials** | 11471 | 11509 | 11502 |

**Table 6.** GAMLSS Model selection based on BIC: admission stroke severity centile for ICH.

| **Distributions**<br>**Additive terms** | **Negative binomial** | **Beta binomial** | **Delaporte** |
| --- | --- | --- | --- |
| **Linear** | **1879** | 1890 | 1882 |
| **Cubic splines** | 1898 | 1925 | 1913 |
| **Fractional polynomials** | 1901 | 1913 | 1909 |

**Table 7.** Eligibility criteria and method selection.

|  | **Method** | **Robust**<br>**Model fit**<br>**Assessment**<br>**framework** | **If often used**<br>**in applied**<br>**medical**<br>**research** | **Clear**<br>**documentation of**<br>**coding in a**<br>**statistical**<br>**software** |
| --- | --- | --- | --- | --- |
| **Parametric** | **GAMLSS** | √ | √ | √ |
|  | S-distribution <sup>45</sup> | x | x | x |
|  | Thompson and Theron <sup>46</sup> | √ | x | x |
| **Semi-parametric** | **Quantile regression, estimate separately <sup>21</sup>** | √ | √ | √ |
|  | Quantile regression, estimate together <sup>47</sup> | x | x | x |
| **Non-parametric** | **HRY** | x | √ | √ |
|  | Adapted HRY <sup>48</sup> | x | x | √ |
|  | Kernel density <sup>49</sup> | x | x | x |
|  | Non-Gaussian quantile <sup>50</sup> | x | x | x |

### Appendix C Parameters estimated in GAMLSS model

In Figure 8 for IS patients, the estimated mean, dispersion and skewness parameters are consistent across three fitted distributions under the assumption of linear additive terms. Except for the negative binomial model with no skewness parameter, the mean parameter is monotonically decreasing over time. The dispersion and skewness parameters increased over time. We obtained similar results in cubic spline models; the only difference is that the dispersion parameter increased initially in the first six hours and then decreased. However, in fractional polynomial models, the mean parameter decreased in the first six hours and then increased. The dispersion parameter increased dramatically in the first two hours, peaked at four hours and gradually decreased when fitting negative binomial distribution. The dispersion parameter increased in the first six hours and decreased in the following hours when fitting beta-binomial and Delaporate distributions. The skewness also increased in the first six hours and decreased afterwards when fitting the beta-binomial distribution. But the skewness parameter is very different when fitting the Delaporate distribution, which dropped dramatically after an hour and remained unchanged over time.

**Figure 8.**
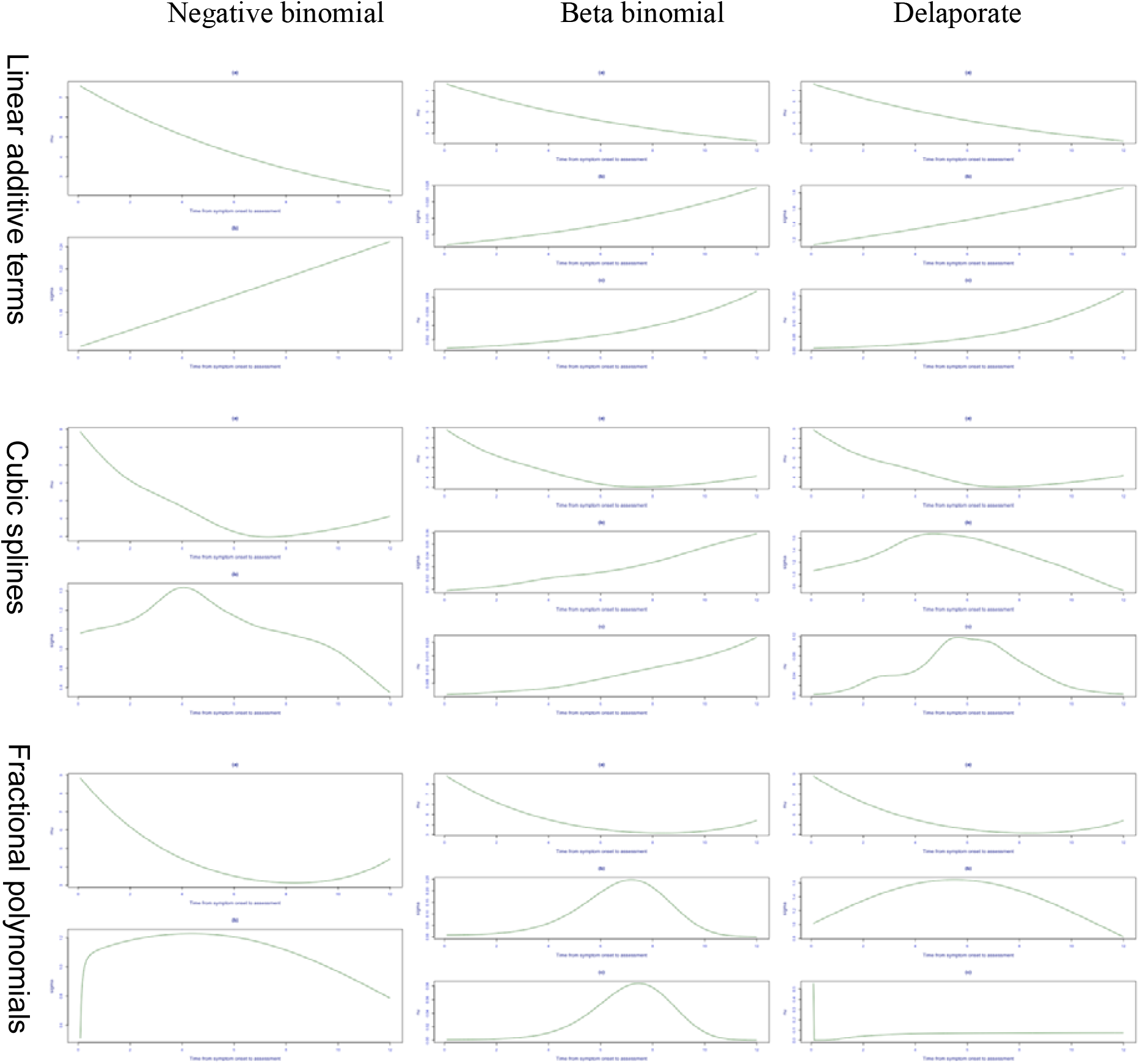
Estimated parameters from GAMLSS model: admission stroke severity centile for IS patients

When modelling the ICH stroke severity centiles, we obtain a consistent pattern for the estimated parameters in Figure 9, except for fractional polynomial models. The inconsistent estimated parameters observed in models with nonlinear additive terms may be attributed to outliers or extreme values that are not averaged over time.

**Figure 9.**
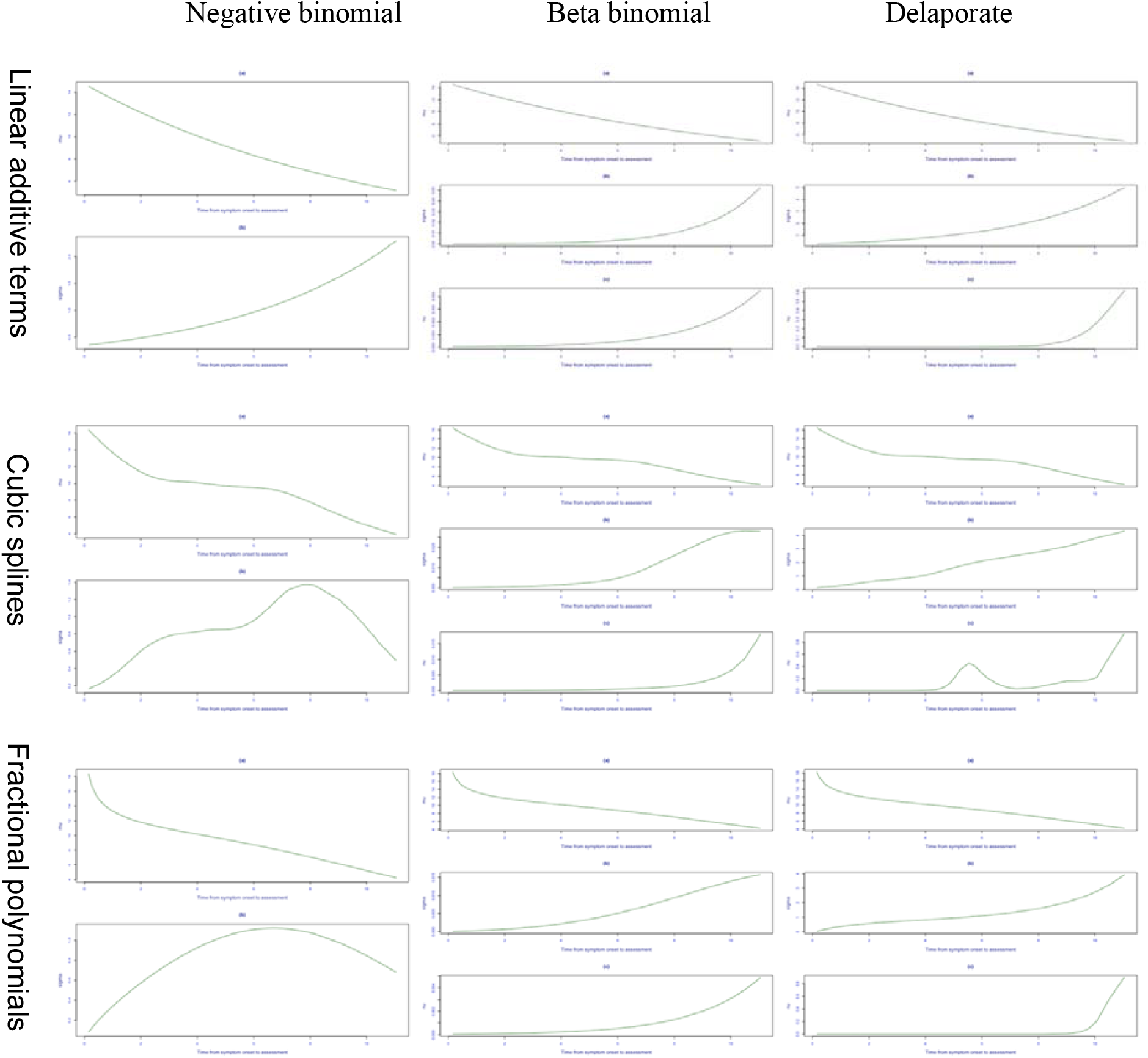
Estimated parameters from GAMLSS model: admission stroke severity centile for ICH patients

### Appendix D Crude centiles calculated in the HRY method

The comparison between Figures 10 and 11 highlights the impact of selecting different values for *k* (proportion of observations used in the sliding window) in the HRY method’s first step when estimating raw centiles.

**Figure 10.**
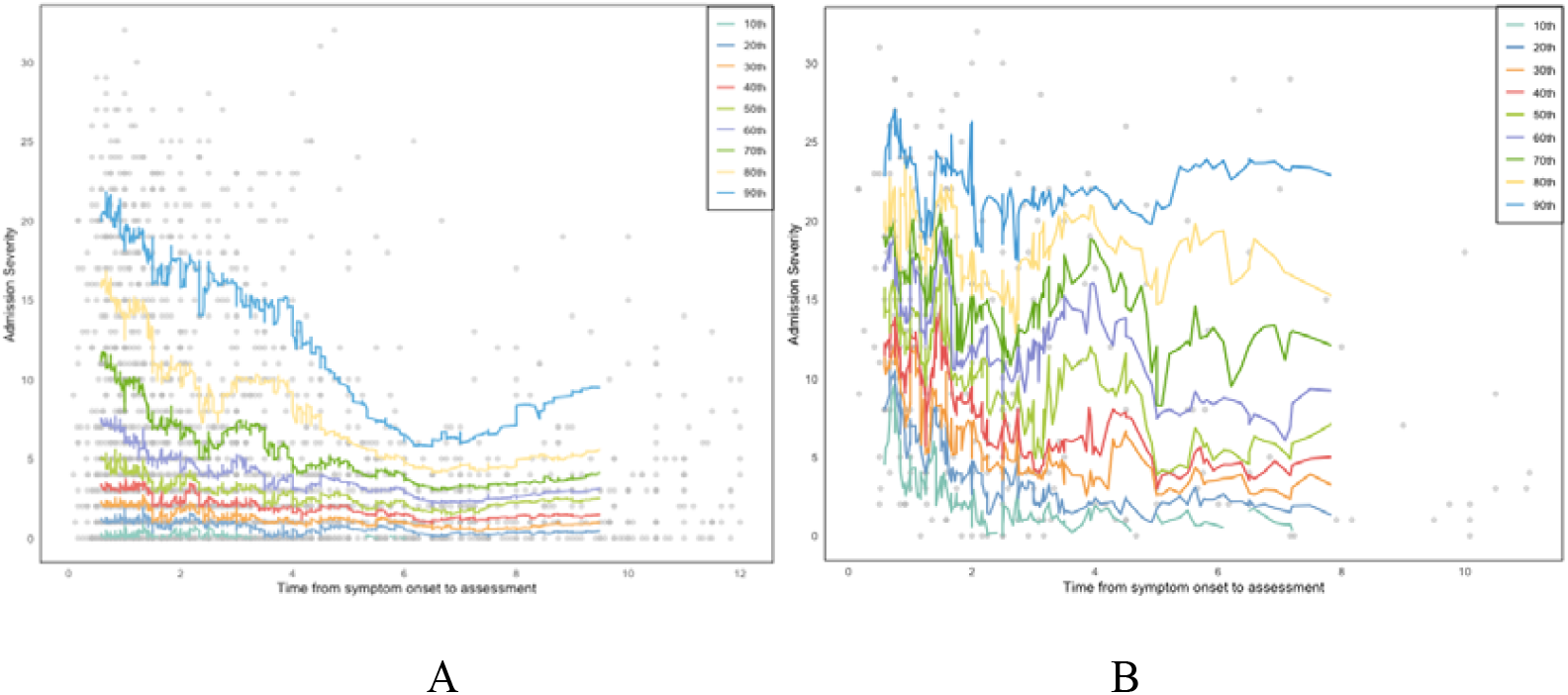
Raw centiles calculated in the HRY method before smoothing when k=0.1: (A) IS;

**Figure 11.**
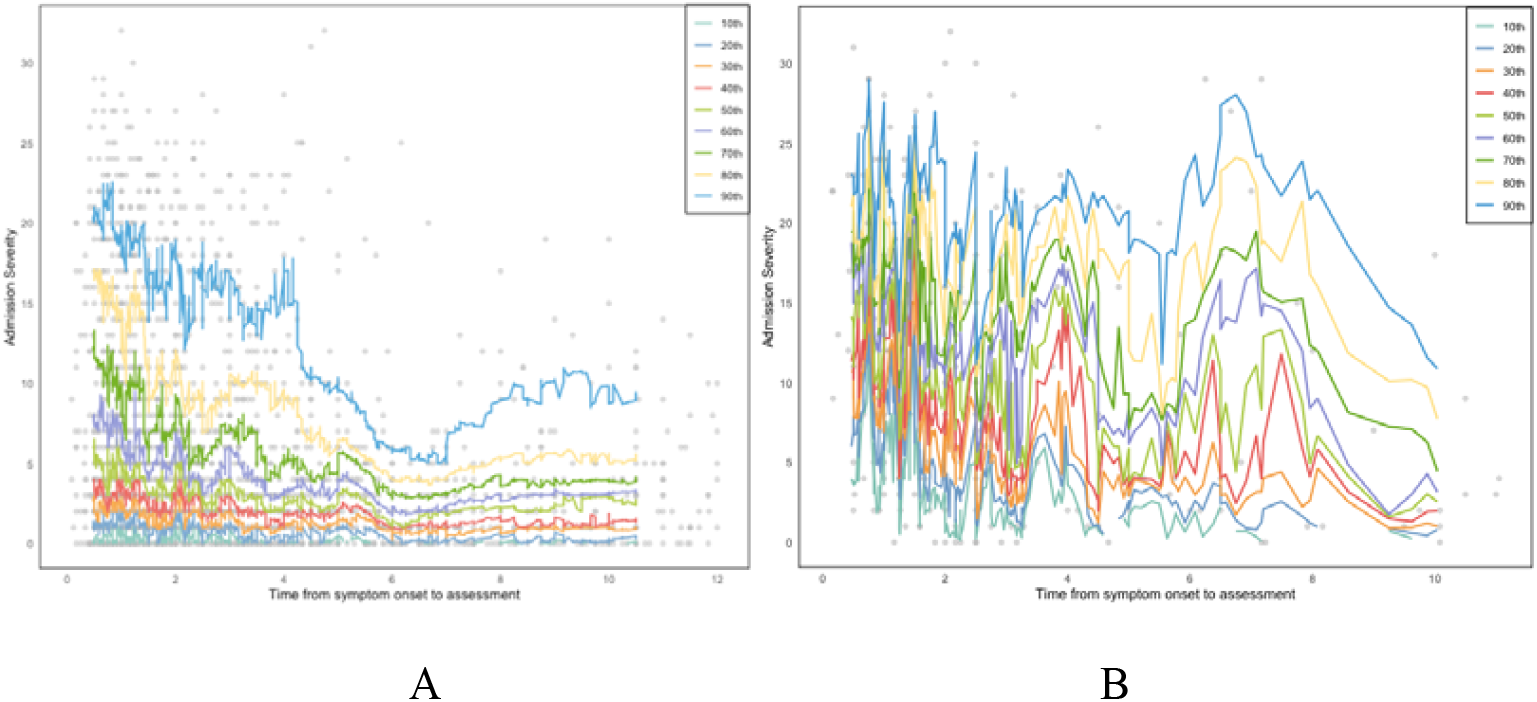
Raw centiles calculated in the HRY method before smoothing when k=0.05: (A) IS;

**Figure 12.**
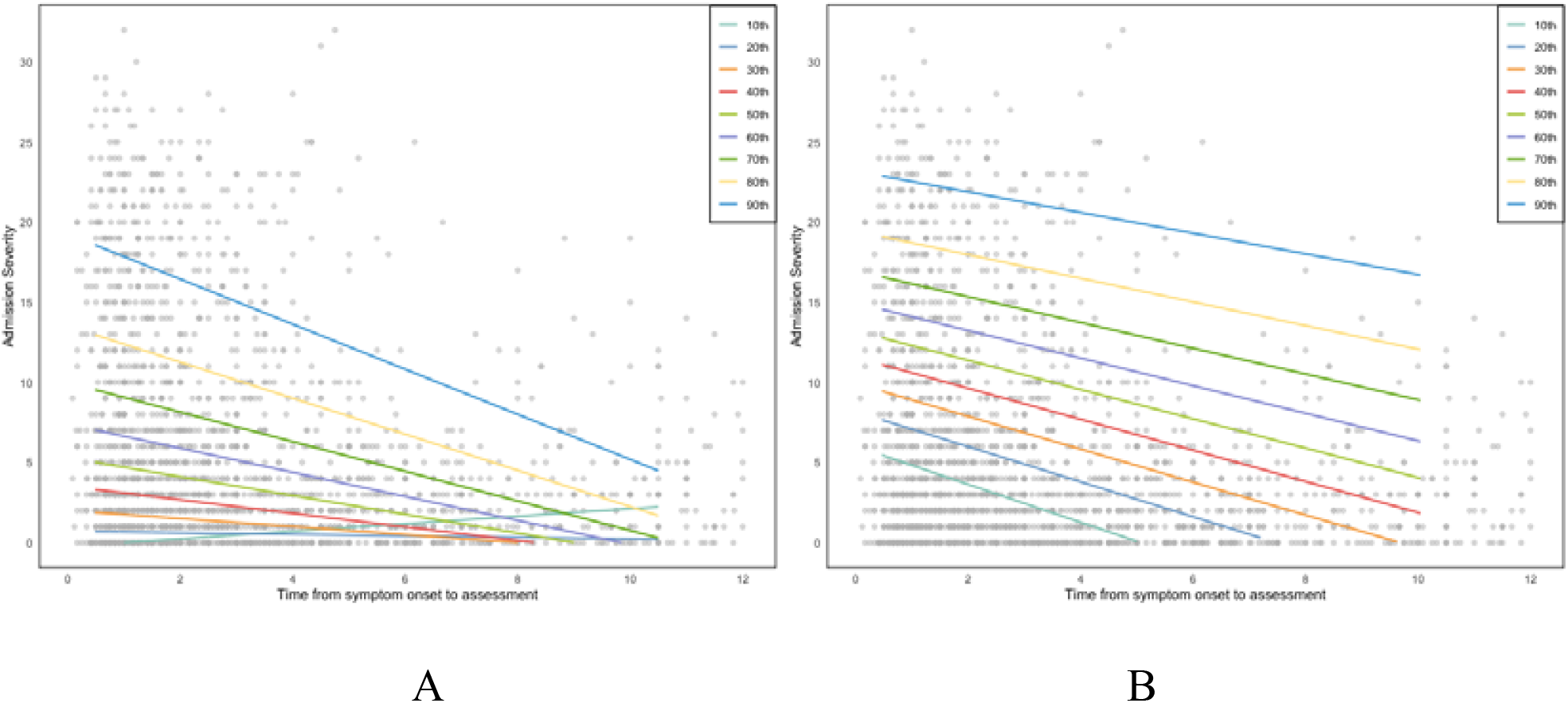
k=0.05, p=1, q_0=2: (A) IS; (B) ICH

**Figure 13.**
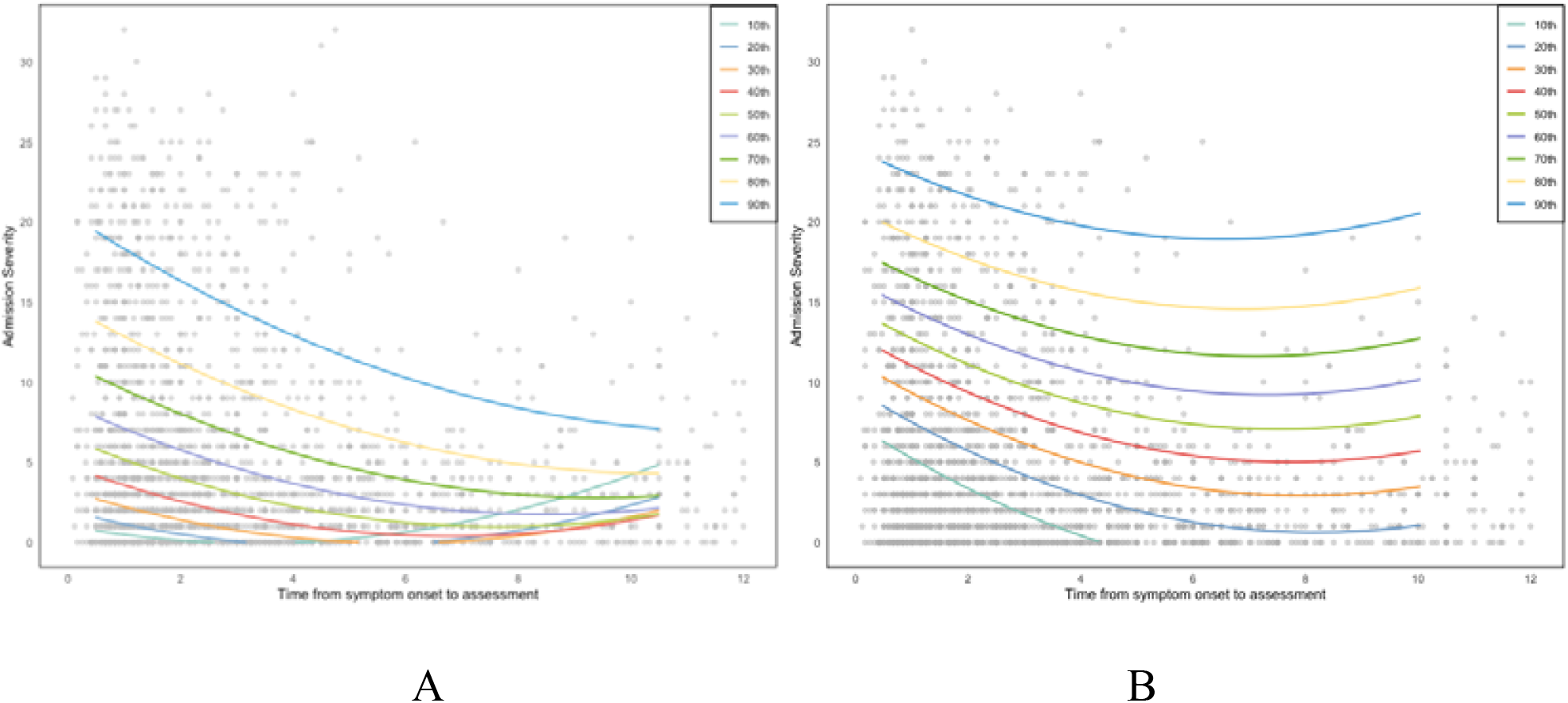
k=0.05, p=2, q_0=2: (A) IS; (B) ICH

**Figure 14.**
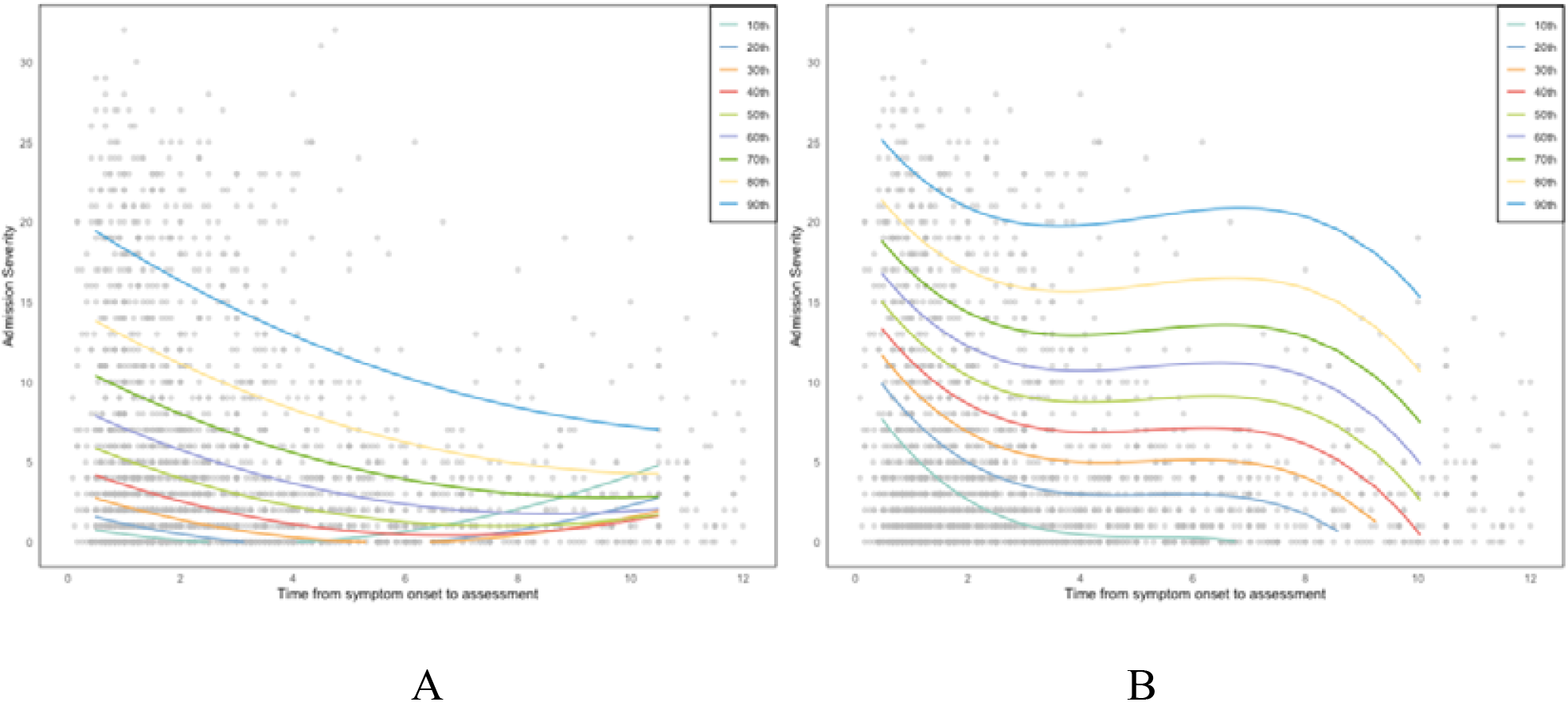
k=0.05, p=3, q_0=2: (A) IS; (B) ICH

**Figure 15.**
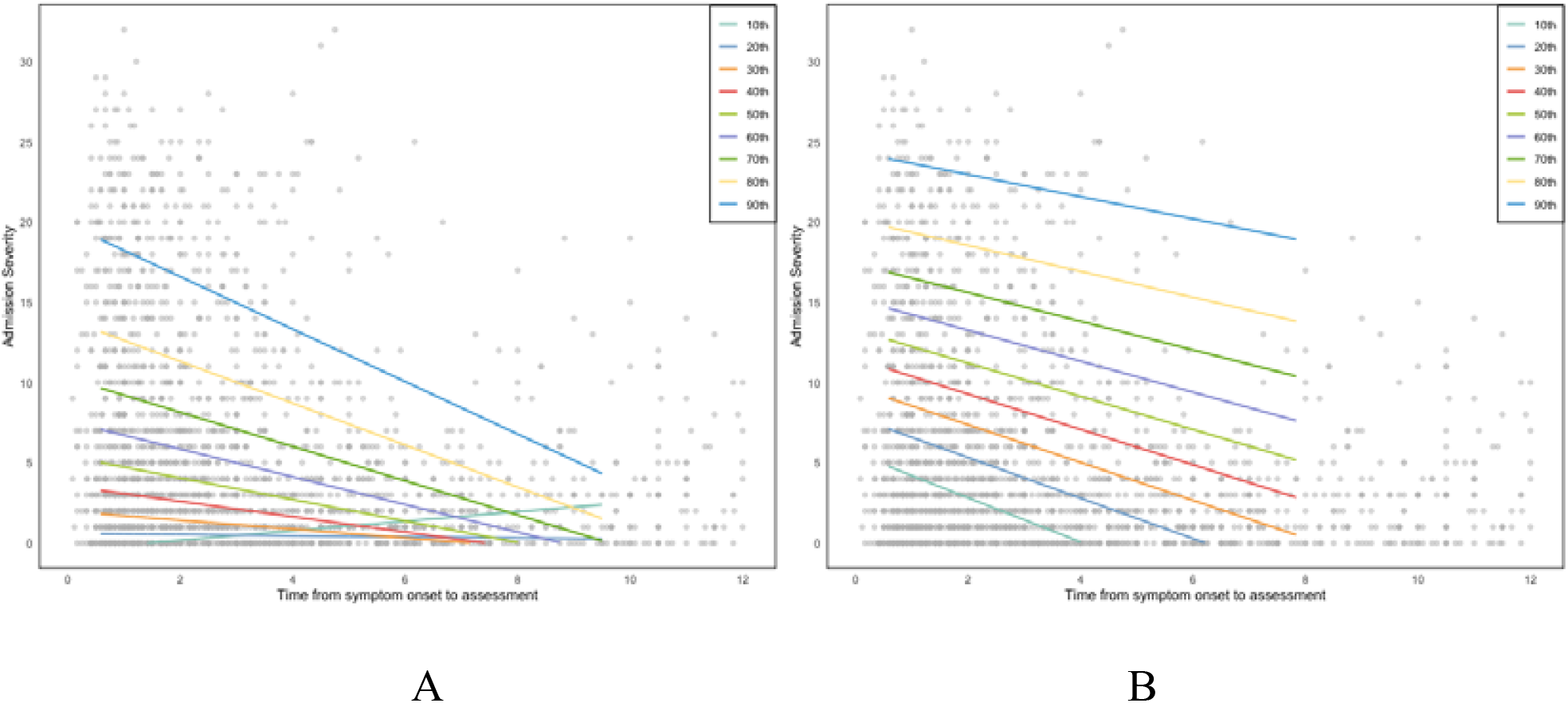
k=0.1, p=1, q_0=2: (A) IS; (B) ICH

**Figure 16.**
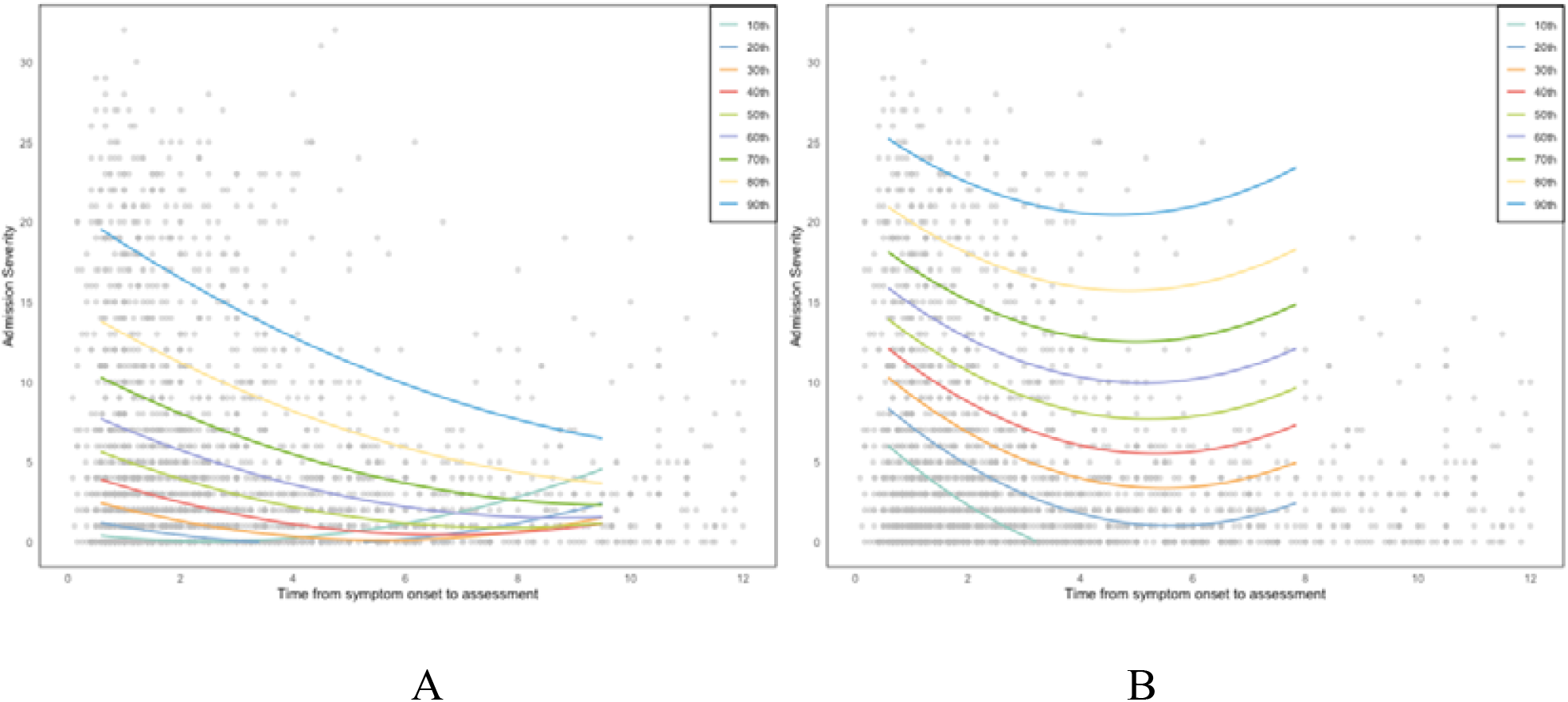
k=0.1, p=2, q_0=2: (A) IS; (B) ICH

**Figure 17.**
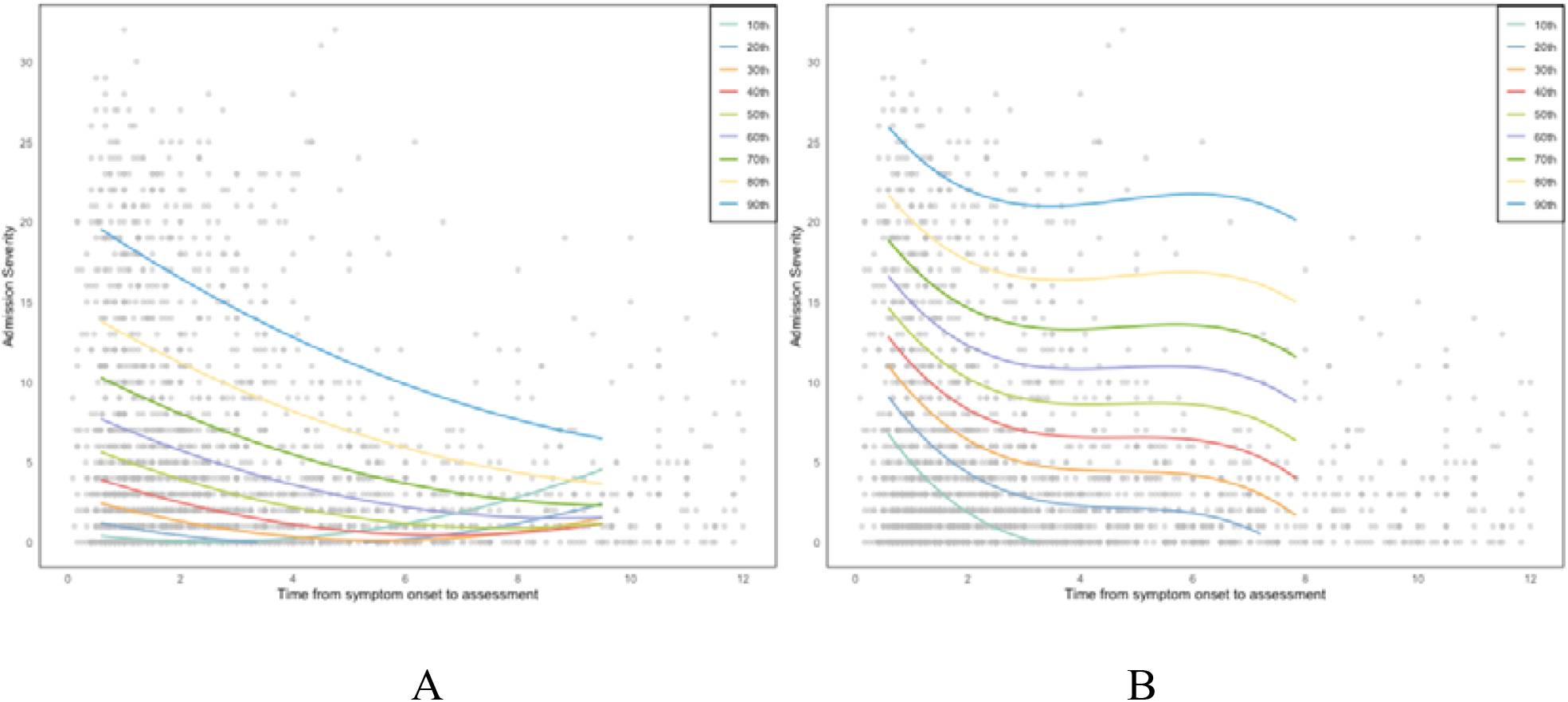
k=0.1, p=3, q_0=2: (A) IS; (B) ICH

With *k*= 0.1 (Figure 10), the centile curves are smoother and less variable, as the larger window incorporates more data points, reducing the influence of outliers and local fluctuations. For ischaemic stroke patients (A), the centiles remain relatively consistent, while for ICH patients (B), the variability is slightly more noticeable but still moderate. In contrast, with *k* = 0.05 (Figure 11), the smaller window introduces greater sensitivity to local data characteristics, resulting in noisier and more irregular centile curves, particularly for lower percentiles (e.g., 10th and 20th). This effect is especially pronounced in ischaemic stroke data due to the larger proportion of zero values amplifying the variability.

Overall, the selection of k is quite arbitrary in the recommended range. A larger *k* provides a better balance between local adaptation and smoothing, minimising variability while preserving the overall shape of the centile curves. On the other hand, a smaller k omits fewer data points from two ends and provides a more complete centile estimation for both IS and ICH data. The selection of k is a trade-off between smoothness and completeness of the estimated centile curves. Therefore, we made *k*= 0.1 a preferable choice for generating more reliable and stable raw centile estimates prior to the smoothing step with the compromise of omitting slightly more data points at the two ends.

The choice of *k* somewhat arbitrary within the recommended range and involves a trade-off between smoothness and completeness of the estimated centile curves. A larger *k* offers a better balance between local adaptation and smoothing, reducing variability while preserving the overall shape of the centile curves. Conversely, a smaller retains more data points at both ends, leading to a more complete centile estimation. To optimise reliability and stability before the smoothing step, we selected, acknowledging a slight compromise in omitting more data points at the extremes.

### Appendix E HRY smoothed centile curves from different sets of parameters

The results in Appendix E demonstrate how the choice of a difference set of parameters influences the estimated centile curves in the HRY method.

Focusing on *p*, the degree of the polynomial for smoothing, we observe that *p*= 1 imposes overly linear trends, failing to capture the curvature inherent in the stroke severity. Conversely, *p*= 3 introduces excessive flexibility, occasionally creating artifacts in sparse regions, particularly in later time intervals. The quadratic smoothing polynomial (*p*= 2) achieves a balance between flexibility and stability, effectively capturing non-linear trends while preserving smooth transitions across time.

Overall, *k*= 0.1, *p*= 2, and q_0_ =2 emerge as the preferred parameters for constructing stroke severity centiles. This choice balances smoothness and adaptability, effectively handling the sparse data and variable distributions typical of stroke severity on admission.

The larger window size (*k*= 0.1) provides more robust local regression estimates, reducing variability and irregularities, while the quadratic smoothing polynomial (*p*= 2) captures nonlinear trends without overfitting. Additionally, q_0_ =2 accounts for skewness in centile intervals, ensuring a better fit across different percentiles. However, it is important to note that this preference is based on informal justification, as no formal model diagnostic tool or model selection procedure is designed explicitly for the HRY method. The choice of parameters relies on subjective evaluation of smoothness and interpretability rather than objective statistical criteria.

### Appendix F Eligibility criteria and method selection

The original article reviewed 30 methods ^16^. After excluding 21 methods that either treated time as a categorical variable (grouping methods) or required a normal distribution or normality transformation of the response variable, nine methods remained for further evaluation based on additional eligibility criteria (Table 7). Methods marked with “√ “ were deemed eligible, while those marked with “x” were not. From this selection, we identified GAMLSS, Quantile Regression (estimated separately), and the HRY method as the most suitable approaches for modelling admission stroke severity centiles.

